# Combining family history, polygenic scores, and childhood trauma to predict major depressive disorder

**DOI:** 10.64898/2026.11.19.26368522

**Authors:** Rujia Wang, Helena L. Davies, Sang-Hyuck Lee, Johan Zvrskovec, Christopher Hübel, Saakshi Kakar, Chelsea Malouf, Laura Meldrum, Yuhao Lin, Iona Smith, Gursharan Kalsi, Henry C. Rogers, Shannon Bristow, Molly R. Davies, Donald M. Lyall, Allan H. Young, Anthony J. Cleare, Katrina A. S. Davis, Roland Zahn, Victor A. Gault, Le Roy C. Dowey, Ruth K. Price, Keith G. Thomas, James T. R. Walters, Daniel J. Smith, Chérie Armour, Ian R. Jones, Nathalie Kingston, John R. Bradley, Andrew M. McIntosh, the NIHR BioResource consortium, the GLAD Study, Matthew Hotopf, Jonathan R.I. Coleman, Evangelos Vassos, Thalia C. Eley, Raquel Iniesta, Gerome Breen

## Abstract

**Objective:** Risk stratification for major depressive disorder (MDD) remains limited by clinical heterogeneity and modest genomic prediction. We examined whether integrating family history, polygenic risk scores (PRS), childhood trauma, and background factors improves prediction of lifetime MDD diagnosis and its severity, relative to models including only subsets of these predictors.

**Methods:** Analyses included 9,927 cases and 4,452 controls from the Genetic Links to Anxiety and Depression Study and NIHR BioResource (GLAD+) and 40,667 cases and 70,755 controls from UK Biobank (UKB). Lifetime MDD was defined using DSM-5 based online assessments. Predictors included family history of multiple psychiatric disorders (mFH), multi-trait PRSs (mPRS), childhood trauma, and background factors (sex and ten genetic principal components). Nested elastic net regression and XGBoost were used to predict MDD within and across cohorts.

**Results:** Multivariable models integrating mFH, mPRS, childhood trauma, and background factors demonstrated strong risk classification (AUC=0.83 in GLAD+; 0.74 in UKB). Family history consistently outperformed multi-trait PRS (GLAD+: 0.74 vs. 0.66; UKB: 0.65 vs. 0.60). External validation showed preserved classification (AUC=0.71-0.82), with attenuation reflecting differences in case severity across cohorts. Models explained up to 27% of the variance in the number of depressive episodes.

**Conclusions:** Multivariable integration of familial, environmental, and genetic factors yields robust and transferable prediction of MDD. Family history and childhood trauma consistently outperform current PRS for both case classification and severity prediction, underscoring the continued importance of integrating clinical data into prediction models in the genomic era.

## Introduction

Major depressive disorder (MDD) is one of the most common psychiatric disorders worldwide, with a lifetime prevalence of 15-18%(1), and 35-75% of individuals with a first episode go on to have recurrences(2). MDD impairs social and occupational functioning, reduces health-related quality of life, and elevates suicide risk(3, 4). Many biological, environmental, and sociodemographic risk factors have been identified(5, 6), and clinicians already weigh family history, early adversity, and demographic factors informally when judging risk and prognosis. Yet reliable tools that combine these factors for individualized risk stratification remain limited(7, 8).

Family history of depression is a consistently robust predictor. The offspring of those with MDD are two-to-three times more likely to develop MDD(5), and family history of other psychiatric disorders, including anxiety disorders and bipolar disorder, is also predictive(6). Family history reflects a combination of common and rare genetic effects together with familial environments and exposures(6, 9). Its accuracy, however, depends on the number and age of relatives, the reporter’s knowledge, and whether relatives have received or disclosed a diagnosis; stigma and historical under-diagnosis mean that familial risk is unknown or incomplete for many individuals(10).

Genome-wide association studies (GWASs) have identified over 600 loci associated with depression(11), and polygenic risk scores (PRSs) for depression now explain a modest but increasing proportion of variance (5-6%)(11). PRSs capture only the common-variant component of inherited risk, and integrating them into clinical prediction remains a difficult challenge in psychiatry(12). One approach is multi-polygenic scoring (mPRS), in which PRSs for several genetically correlated disorders and traits are combined using supervised machine learning(13–15). Because PRSs do not depend on relatives’ diagnoses being known, they may add most where family history is unavailable(10).

Childhood trauma and sex are also well-established risk factors. Childhood trauma is associated with increased risk of depression(16–18) and with earlier onset, greater chronicity, and treatment-resistant depression(17). MDD occurs approximately twice as often in females as in males(19).

We tested whether prediction of MDD could be improved by integrating family history of multiple psychiatric disorders with mPRS, childhood trauma, and sex. We used supervised machine learning, which optimises predictive accuracy from multivariable data without requiring prespecified assumptions about causal relationships between predictors and outcomes(14, 20); elastic net regression handles correlated predictors and permits comparison across models using a common predictor set. We used two large UK cohorts, a clinically enriched sample (the Genetic Links to Anxiety and Depression Study and other NIHR BioResource cohorts; GLAD+) and a population-based sample (UK Biobank; UKB), training and testing models within each cohort and validating them externally in the other. We examined: (1) the relative and incremental contributions of family history, mPRS, childhood trauma, and background factors to classification of lifetime MDD and its severity; and (2) the generalizability and calibration of prediction across cohorts.

## Methods

### Study sample

GLAD+ combines two UK cohorts: the NIHR BioResource Genetic Links to Anxiety and Depression (GLAD) Study (https://www.gladstudy.org.uk)(21), which recruits participants with lifetime depression and/or anxiety, and healthy volunteers from other NIHR BioResource (https://bioresource.nihr.ac.uk/) cohorts. Both groups were recontacted during the COVID-19 pandemic for the COPING Study, through which the measures used here were collected. Participants provide demographic, environmental, and genetic data and consent to medical record linkage and recontact (Supplementary Methods).

UKB is a population-based cohort of over 500,000 UK residents aged 40-69 years at recruitment (2006-2010). Online Mental Health Questionnaires were completed in 2016-17 (MHQ1; over 157,000 respondents)(22) and 2022 (MHQ2; over 169,000), with more than 206,000 participants completing at least one. Family history of psychiatric disorders among first-degree relatives was collected in MHQ2. All participants provided informed consent.

### Measurements

#### Major depressive disorder (MDD)

Lifetime MDD was assessed in both cohorts with the adapted Composite International Diagnostic Interview-Short Form (CIDI-SF) based on DSM-5 criteria(22). Cases reported at least five symptoms, including depressed mood or loss of interest, nearly every day for at least two weeks. Controls did not meet these criteria and reported neither a diagnosis of depression nor any depressive episode. In UKB, cases met criteria in either MHQ1 or MHQ2 (Table S1). After exclusions for depressive episodes among controls, genotype availability and quality, non-European genetic ancestry, participation in UKB (identified by duplicate genotypes), and missing predictor data, the final GLAD+ sample comprised 9,927 MDD cases and 4,452 controls (Figure S1). UKB included 40,667 cases and 70,755 controls (Table S1).

Three severity measures were available. The number of depressive episodes lasting two or more weeks was reported in categories from “1” to “13 or more” in GLAD+ and as a count in UKB, which was mapped onto the GLAD+ categories (Supplementary Methods). Age of onset was the self-reported age at the first two-week period of depression. In GLAD+ only, the degree of treatment-resistance was assessed as a composite score (0-14) combining the 4-item Maudsley Staging Method(23) and the Patient Health Questionnaire-9(24) (Table S2).

#### Family history (FH)

In GLAD+, participants reported whether any family member had ever been diagnosed with each of 22 psychiatric disorders (Table S3; optional survey). In UKB, participants reported whether any first-degree blood relative had been diagnosed with each of nine disorders (Table 1; MHQ2). Each disorder was coded as a binary indicator of at least one affected relative. Multiple family history (mFH) therefore denotes a set of disorder-specific indicators entered simultaneously into the model, not a count or composite. Main analyses used the nine disorders assessed in both cohorts; the full 22 GLAD+ items were used only in a sensitivity analysis.

**Table 1.** Characteristics of participants in GLAD+ and UKB.

| Characteristics | GLAD+ |  |  | UKB |  |  |
|---|---|---|---|---|---|---|
|  | **Total (n=14,379)** | **MDD cases (n=9,927)** | **Controls (n=4,452)** | **Total (n=111,422)** | **MDD cases (n=40,667)** | **Controls (n=70,755)** |
| **Age** (mean ± sd) | 48.4 ± 15.6 | 44.3 ± 14.7 | 57.5 ± 13.5 | 56.0 ± 7.5 | 54.2 ± 7.4 | 57.0 ± 7.4 |
| **Sex** (female, %) | 10,179 (70.8) | 7,945 (80.0) | 2,234 (50.3) | 62,452 (56.0) | 28,457 (70.0) | 33,995 (48.0) |
| **Family history** (yes, %) |  |  |  |  |  |  |
| depression | 7,116 (49.5) | 6,119 (61.6) | 997 (22.4) | 28,373 (25.5) | 17,352 (42.7) | 11,021 (15.6) |
| anxiety | 4,552 (31.7) | 3,960 (39.9) | 592 (13.3) | 11,248 (10.1) | 6,683 (16.4) | 4,565 (6.5) |
| bipolar disorder | 1,219 (8.5) | 1,094 (11.0) | 125 (2.8) | 3,274 (2.9) | 1,928 (4.7) | 1,346 (1.9) |
| eating disorder | 925 (6.4) | 804 (8.1) | 121 (2.7) | 3,248 (2.9) | 1,811 (4.5) | 1,437 (2.0) |
| SCZ | 596 (4.1) | 528 (5.3) | 68 (1.5) | 1,685 (1.5) | 893 (2.2) | 792 (1.1) |
| personality disorder | 475 (3.3) | 443 (4.5) | 32 (0.72) | 1,398 (1.3) | 940 (2.3) | 458 (0.64) |
| psychosis | 584 (4.1) | 525 (5.3) | 59 (1.3) | 1,679 (1.5) | 967 (2.4) | 712 (1.0) |
| ADHD | 647 (4.5) | 572 (5.8) | 75 (1.7) | 2,125 (1.9) | 1,260 (3.1) | 865 (1.2) |
| ASD | 1,177 (8.2) | 1,021 (10.3) | 156 (3.5) | 3,315 (3.0) | 1,915 (4.7) | 1,400 (2.0) |
| **Childhood trauma** (yes, %) | 5,754 (40.0) | 5,020 (50.6) | 734 (16.5) | 31,051 (27.9) | 17,019 (41.8) | 14,032 (19.8) |
| **MDD PRS** (mean ± sd) | 0 ± 1 | 0.16 ± 0.97 | -0.36 ± 0.98 | 0 ± 1 | -0.002 ± 1 | 0.001 ± 1 |
| **Age of onset** (mean ± sd) |  | 21.3 ± 11.2 |  |  | 33.4 ± 15.0 |  |
| **MDD episodes** (median, IQR) |  | 6 (4-13) |  |  | 2 (1 - 4) |  |

Values are n (%) for categorical variables, mean ± standard deviation (SD) for age, age of onset and MDD PRS, and median (interquartile range, IQR) for the number of MDD episodes. Age of onset and number of episodes are shown for MDD cases only. The number of episodes was recorded in categories from “1” to “13 or more” in GLAD+ and as a count in UKB, which was mapped onto the GLAD+ categories; “13 or more” was coded as 13. PRSs were standardised using the mean and SD of the score in the European 1000 Genomes reference sample, placing scores in both cohorts on the same scale. Family history was reported for any relative in GLAD+ and for first-degree relatives only in UKB. ADHD, attention-deficit/hyperactivity disorder; ASD, autism spectrum disorder; GLAD+, Genetic Links to Anxiety and Depression Study and NIHR BioResource; IQR, interquartile range; MDD, major depressive disorder; PRS, polygenic risk score; SCZ, schizophrenia; SD, standard deviation; UKB, UK Biobank.

#### Polygenic risk scores

PRSs were calculated with the GenoPred pipeline(25) using MegaPRS(26) under the LDAK heritability model, with the best-fitting model selected by pseudovalidation on the GWAS summary statistics, and were standardised to an ancestry-matched reference sample (Supplementary Methods). In both cohorts we generated PRSs for MDD, anxiety, ADHD, bipolar disorder, ASD, PTSD, and BMI from European-ancestry GWASs that excluded UKB samples (Table S4). Following the multi-polygenic score approach(15), these seven PRSs were entered as separate standardised predictors rather than combined into a single score. PRSs for 15 further traits, several derived from GWASs that include UKB participants, were generated in GLAD+ and used only in a GLAD+ sensitivity analysis (Table S4).

#### Childhood trauma (ChT)

Childhood trauma was assessed in both cohorts with the five-item Childhood Trauma Screener(27), covering emotional, physical, and sexual abuse and emotional and physical neglect; participants reporting any trauma type were classified as exposed (Table S5). Childhood trauma was the environmental exposure selected because it is robustly replicated, was measured identically in both cohorts, and precedes the typical age of onset; adult exposures were either not harmonised across cohorts or were measured after onset.

### Analytical approach

MDD status was predicted using multivariable nested elastic net regression(28), which mitigates overfitting and handles multicollinearity(15). Nested cross-validation used five randomly assigned outer folds for model evaluation and ten inner folds, stratified by case status, for tuning α and λ by grid search (caret default grid) on classification accuracy (binary outcomes) or root mean squared error (continuous outcomes) (Figure 1). Performance was quantified by the area under the receiver operating characteristic curve (AUC), computed from pooled out-of-fold predictions across the outer folds, and by mean squared error for continuous outcomes. AUCs of predictor groups were compared using DeLong’s test for paired ROC curves on identical outer folds, with FDR and Bonferroni correction; because DeLong’s test may be miscalibrated for nested models(29), comparisons with the full model were interpreted alongside the magnitude of the AUC difference. Class weights adjusted for case-control imbalance (Supplementary Methods).

**Figure 1.**
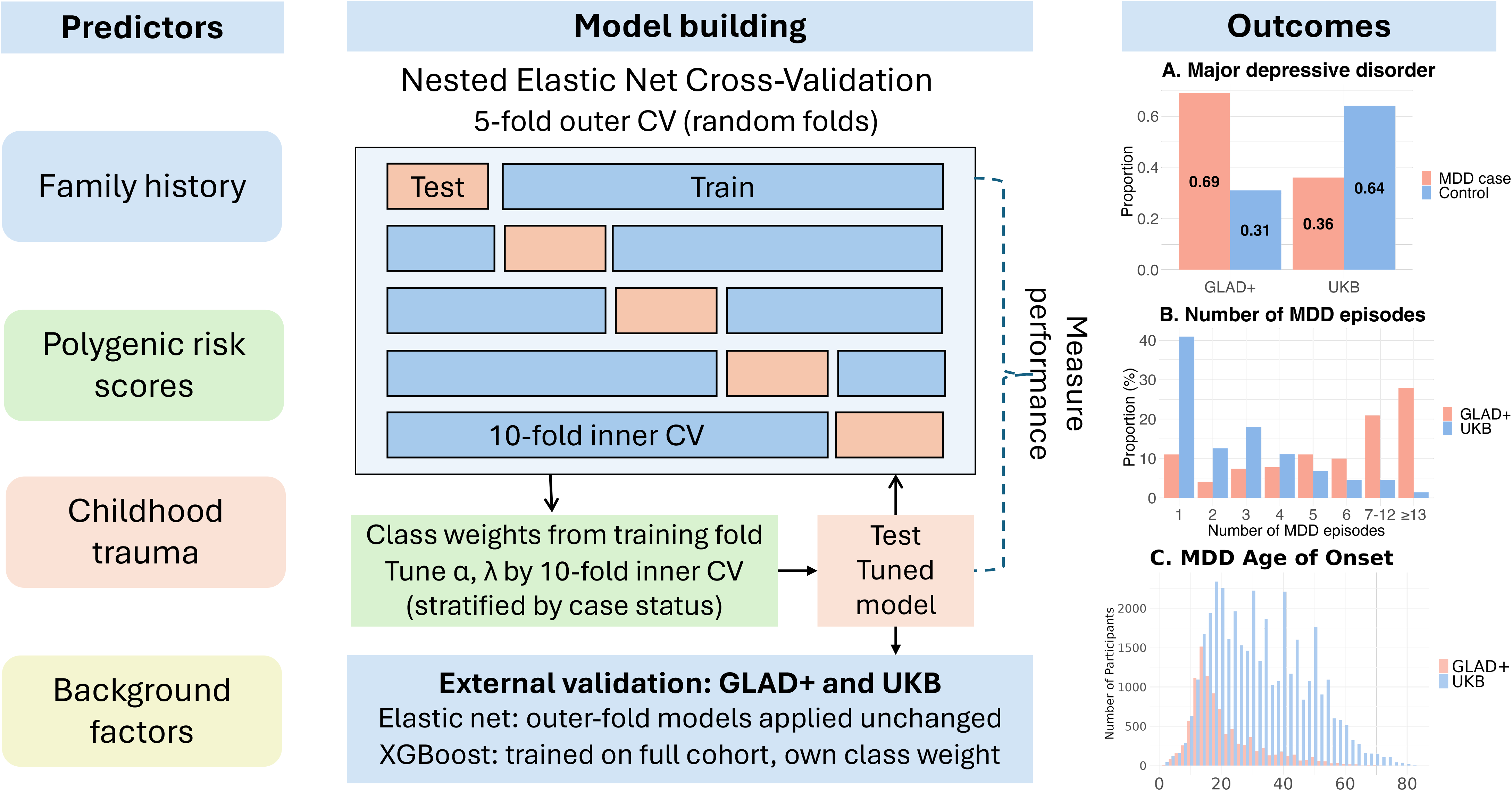
Flow chart of prediction model. Predictors were family history of nine psychiatric disorders (mFH), polygenic risk scores for seven psychiatric disorders and related traits (mPRS), childhood trauma, and background factors (sex and ten genetic principal components). Outcomes were MDD diagnosis, number of episodes, and age of onset. Elastic net models were evaluated with nested cross-validation: participants were randomly assigned to five outer folds, each serving in turn as the test set, and α and λ were tuned within each outer training set using ten inner folds stratified by case status. Class weights were computed within each training fold and applied only to the training loss. For external validation, the outer-fold elastic net models trained in one cohort (GLAD+ or UKB) were applied unchanged to the other cohort, and vice versa. XGBoost, used as a sensitivity analysis, was evaluated within each cohort by 5-fold cross-validation stratified by case status and, for external validation, was trained on the full development cohort with a class weight computed from that cohort alone and applied unchanged to the other cohort. MDD, major depressive disorder; UKB, UK Biobank. Alt text: Flow chart of the analysis. Four predictor groups (family history of nine psychiatric disorders, polygenic scores for seven traits, childhood trauma, and sex with ten genetic principal components) enter elastic net models fitted with nested cross-validation (five outer folds for evaluation, ten inner folds for tuning). Models predict MDD diagnosis, number of episodes, and age of onset, are trained in one cohort (GLAD+ or UK Biobank), and are validated in the other.

As a sensitivity analysis for non-linear effects, we applied eXtreme gradient boosting (XGBoost) with the same predictors. A planned hyperparameter grid search did not complete, so prespecified hyperparameters were used (Table S6). XGBoost was evaluated within each cohort by stratified 5-fold cross-validation and, for external validation, trained and class-weighted on one full cohort and applied unchanged to the other (Supplementary Methods).

Predictors were grouped into four domains: (1) mFH for nine psychiatric disorders, (2) mPRS for seven disorders and traits, (3) childhood trauma, and (4) background factors (sex and ten genetic principal components), all entered as penalized predictors in the elastic net and as standard features in XGBoost. Age was not included (see Limitations). Predictors were harmonised across GLAD+ and UKB (Table 1), and analyses used complete cases (Supplementary Methods; Figure S1). Predictor contributions were summarised as standardised coefficients (coefficient × predictor SD), averaged across the five outer-fold models, and as the number of outer folds in which each predictor had a non-zero coefficient.

Models predicting MDD diagnosis, number of depressive episodes, and age of onset were trained within each cohort and evaluated in the other. For external validation, elastic net models trained within the outer folds (80% of the training cohort) were applied to the other cohort; GLAD+-trained outer-fold coefficients were compared with those of a model refit on the full GLAD+ sample (Tables S7-S7a). As a sensitivity analysis of case-mix effects, UKB MDD cases were resampled to match the GLAD+ episode distribution, together with randomly selected controls (n = 2,442, 1,597 cases and 845 controls), with approximate 95% confidence intervals for AUC and R² derived from the estimates and sample sizes(30). We also examined whether the model trained for MDD diagnosis predicted the three severity measures (treatment-resistance in GLAD+ only).

Observed-scale R² was converted to the liability scale(31) assuming a population prevalence of 5% in GLAD+, reflecting its clinically enriched, recurrent case definition, and 15% in UKB, reflecting lifetime MDD in the general population(1, 32); because this conversion depends on the assumed prevalence, estimates across 5-20% are given in Table S8 alongside observed-scale R² and AUC. Positive and negative predictive values (PPV, NPV), sensitivity, and specificity were calculated at prediction-score cut-offs of 0.3-0.7, and calibration plots compared predicted probabilities with observed outcomes.

## Results

GLAD+ cases were more severe: 89% had recurrent episodes and 28% reported 13 or more, whereas 41% of UKB cases had a single episode and 72% had three or fewer (Figure 2). Compared with controls (GLAD+ = 4,452; UKB = 70,755), participants with MDD (GLAD+ = 9,927; UKB = 40,667) were younger, more often female, more likely to report a family history of depression and anxiety, and more likely to have experienced childhood trauma (Table 1).

**Figure 2.**
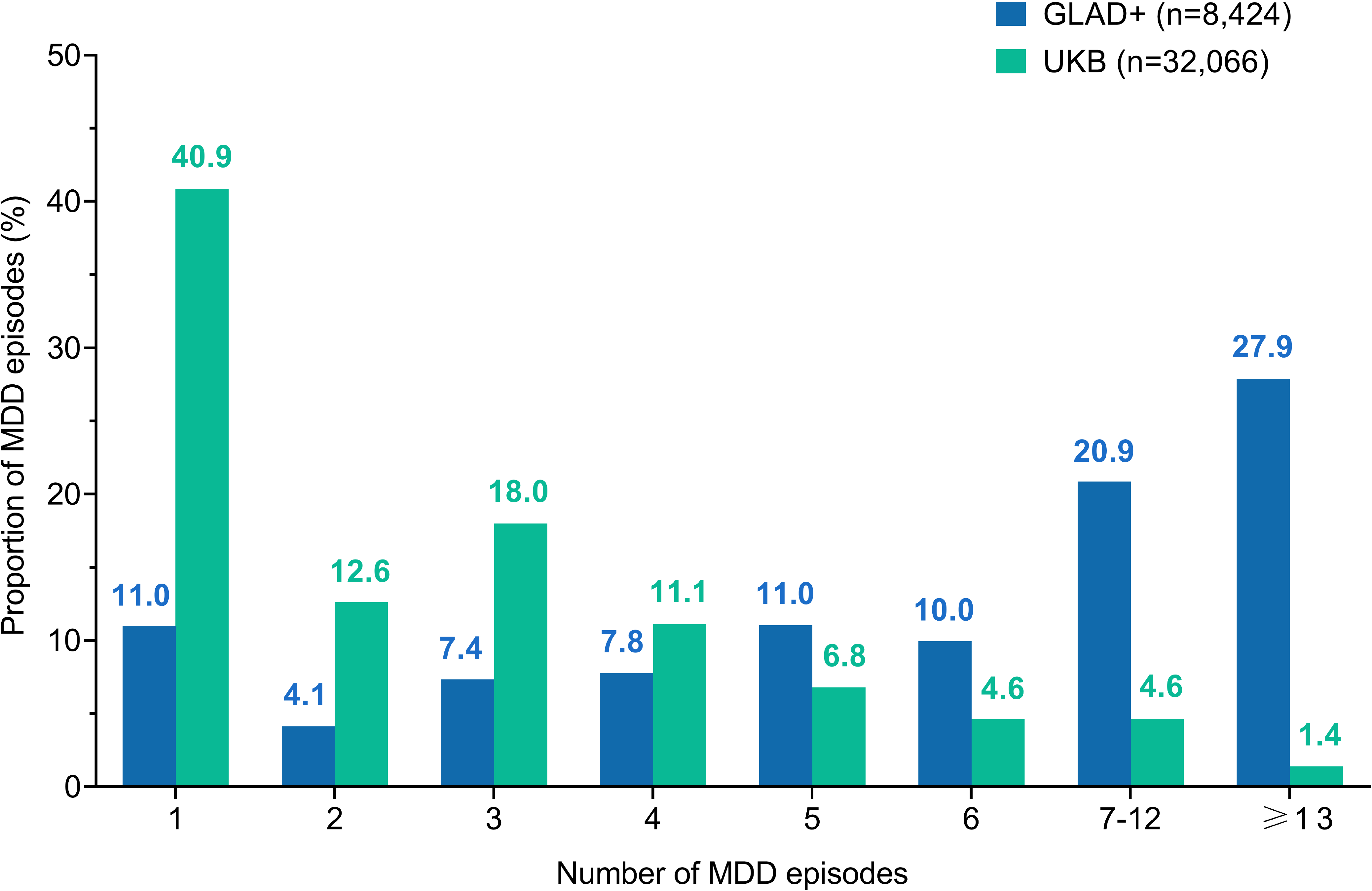
The number of MDD episodes in the GLAD+ and UK Biobank samples included in the prediction analyses. Alt text: Chart comparing the distribution of lifetime depressive episodes among MDD cases in the two cohorts. GLAD+ cases mostly had recurrent depression (89%), and 28% reported 13 or more episodes. In UK Biobank, 41% of cases had a single episode and 72% had three or fewer.

### Prediction of MDD status

In GLAD+, the model integrating family history, mPRS, childhood trauma, and background factors achieved an AUC of 0.83 under nested cross-validation (Figure 3A). Family history was the strongest single domain (AUC=0.74), whereas mPRS (0.66), childhood trauma (0.67), and background factors (0.67) each showed moderate performance alone. In UKB, discrimination was lower but followed the same pattern (Figure 3B): the combined model achieved an AUC of 0.74, and family history (0.65) outperformed mPRS, childhood trauma, and background factors (0.60-0.62; Table S9). The single-trait MDD PRS performed similarly to the mPRS (AUC 0.65 vs 0.66 in GLAD+; 0.59 vs 0.60 in UKB; Table S10), and excluding mPRS from the full model reduced discrimination only slightly (0.82 vs 0.83 in GLAD+; 0.73 vs 0.74 in UKB; Table S11). Each domain improved discrimination beyond background factors alone, with the largest gain from family history (Table S12). Childhood trauma, family history of depression, female sex, MDD PRS, and family history of anxiety had the largest standardised coefficients and were retained in all outer folds in both cohorts (Figure S2). Extending the GLAD+ predictors from the harmonised set to all 22 family history items and 22 PRSs yielded only small gains (full model AUC 0.836 vs 0.833; Table S13).

**Figure 3.**
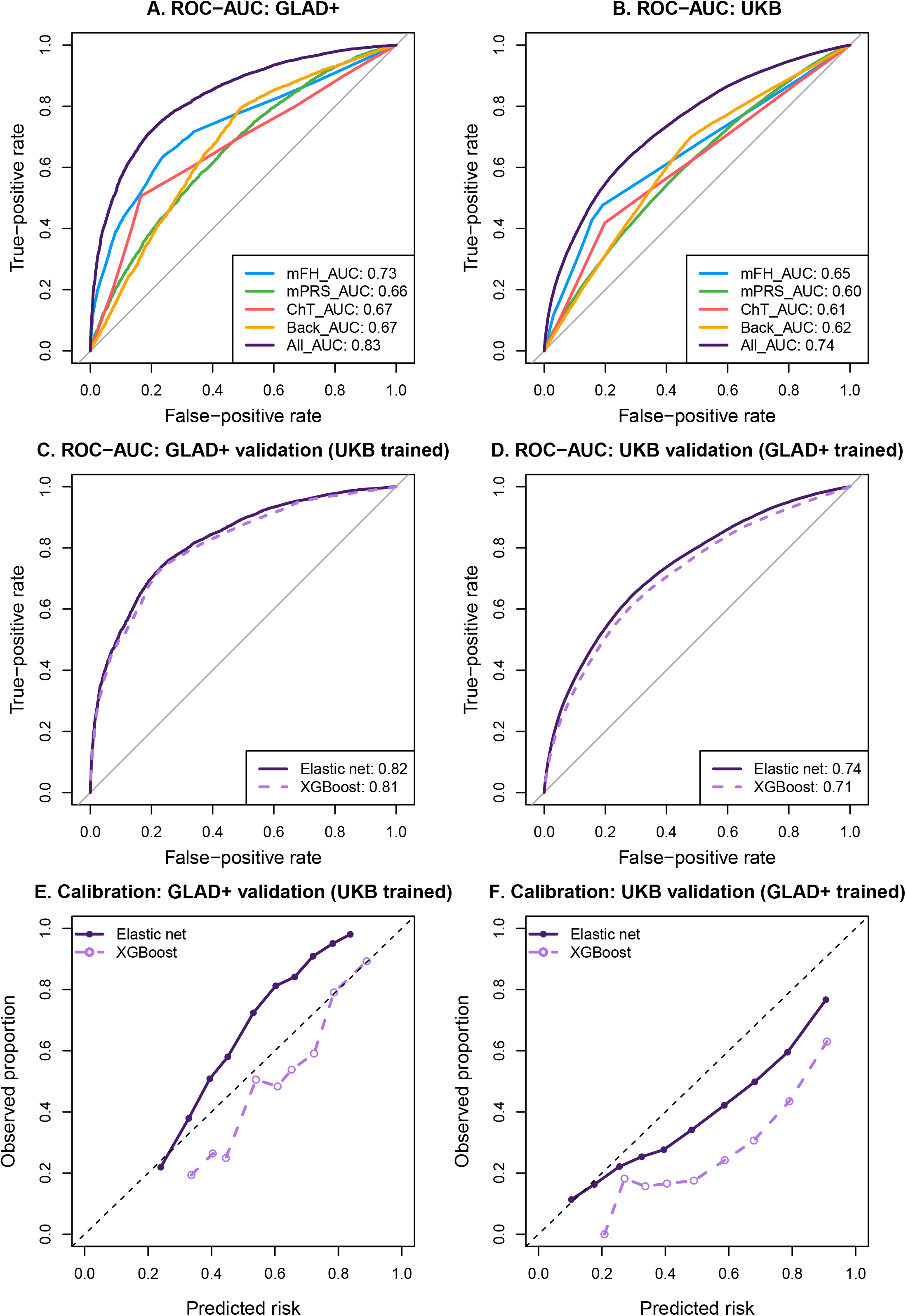
Classification and calibration performance of prediction models for MDD in the GLAD+ and UK Biobank. (A-B) Receiver operating characteristic (ROC) curves showing classification performance of individual predictor domains in GLAD+ (A) and UKB (B) using nested elastic net regression. mFH = family history of multiple psychiatric disorders; mPRS = polygenic risk scores; ChT = childhood trauma; Back = background factors, including sex and ten genetic principal components; All = combined multivariable model including all predictors. (C-D) ROC curves for external validation of elastic net and XGBoost models across cohorts. Panel C shows validation in GLAD+ using models trained in UKB; Panel D shows validation in UKB using models trained in GLAD+. (E-F) Calibration plots for external validation analyses. Observed proportions of MDD are plotted against predicted risk across deciles of predicted probability. The dashed line represents perfect calibration. Solid lines represent elastic net; dashed lines represent XGBoost. Alt text: Six-panel figure. Panels A and B show ROC curves for each predictor group within GLAD+ and UK Biobank; the full model had the highest AUC (0.83 and 0.74), followed by family history (0.74 and 0.65), with polygenic scores, childhood trauma, and background factors lower (0.60-0.67). Panels C and D show external validation ROC curves: UKB-trained models tested in GLAD+ reached AUCs of 0.82 (elastic net) and 0.81 (XGBoost), and GLAD+-trained models tested in UK Biobank reached 0.74 and 0.71. Panels E and F are calibration plots of observed versus predicted risk by decile, showing that UKB-trained models underestimated risk in GLAD+ and GLAD+-trained models overestimated risk at higher predicted probabilities in UK Biobank.

### External validation and calibration

Models transferred across cohorts (Figure 3C-D). UKB-trained models validated in GLAD+ achieved AUCs of 0.82 (elastic net) and 0.81 (XGBoost); GLAD+-trained models validated in UKB achieved 0.74 and 0.71. Models without mPRS performed similarly (elastic net AUC 0.81 in GLAD+; 0.72 in UKB; Table S11). Coefficients of the GLAD+-trained outer-fold models were highly concordant with the full-sample refit (Pearson r = 0.993-0.998; Tables S7-S7a), and liability-scale R² was higher in GLAD+ than in UKB at every assumed prevalence (Table S8).

Calibration differed by learner and setting (Figure 3E-F, Table S14). Elastic net had calibration intercepts closer to zero in all five train-test settings, whereas XGBoost had slopes closer to one and lower Brier scores in four of five. In external validation, both learners departed clearly from ideal calibration: UKB-trained models underestimated risk in GLAD+ (slope 1.40 for both), and GLAD+-trained models overestimated risk at higher predicted probabilities in UKB (slope 0.79 for elastic net and 0.85 for XGBoost).

At a cut-off of 0.5, the UKB-trained model validated in GLAD+ had a PPV of 0.86, sensitivity 0.77, specificity 0.72, and NPV 0.58; the GLAD+-trained model validated in UKB had an NPV of 0.76, specificity 0.79, PPV 0.60, and sensitivity 0.56 (Table S15). These differences largely reflected MDD prevalence (69% in GLAD+ vs 36% in UKB): applying either model’s sensitivity and specificity to the other cohort’s prevalence reproduced its observed predictive values (Table S16). Within-cohort models showed similar patterns (Table S17). The four predictor domains were weakly intercorrelated (r = 0.15-0.17; Table S18).

### Prediction of MDD severity

Applied to severity outcomes (Table S19), the diagnosis-trained model’s predicted risk increased on average across episode categories, although distributions overlapped substantially and the gradient was weaker in UKB (Figure 4). It explained 27% of the variance in the number of episodes in GLAD+ and 11% in UKB, 9% and 6% of the variance in age of onset, and 0.5% of the variance in treatment-resistance (p=1.18x10-5). The diagnosis-trained model therefore carries information relevant to the number of episodes and age of onset, but little for treatment resistance.

**Figure 4.**
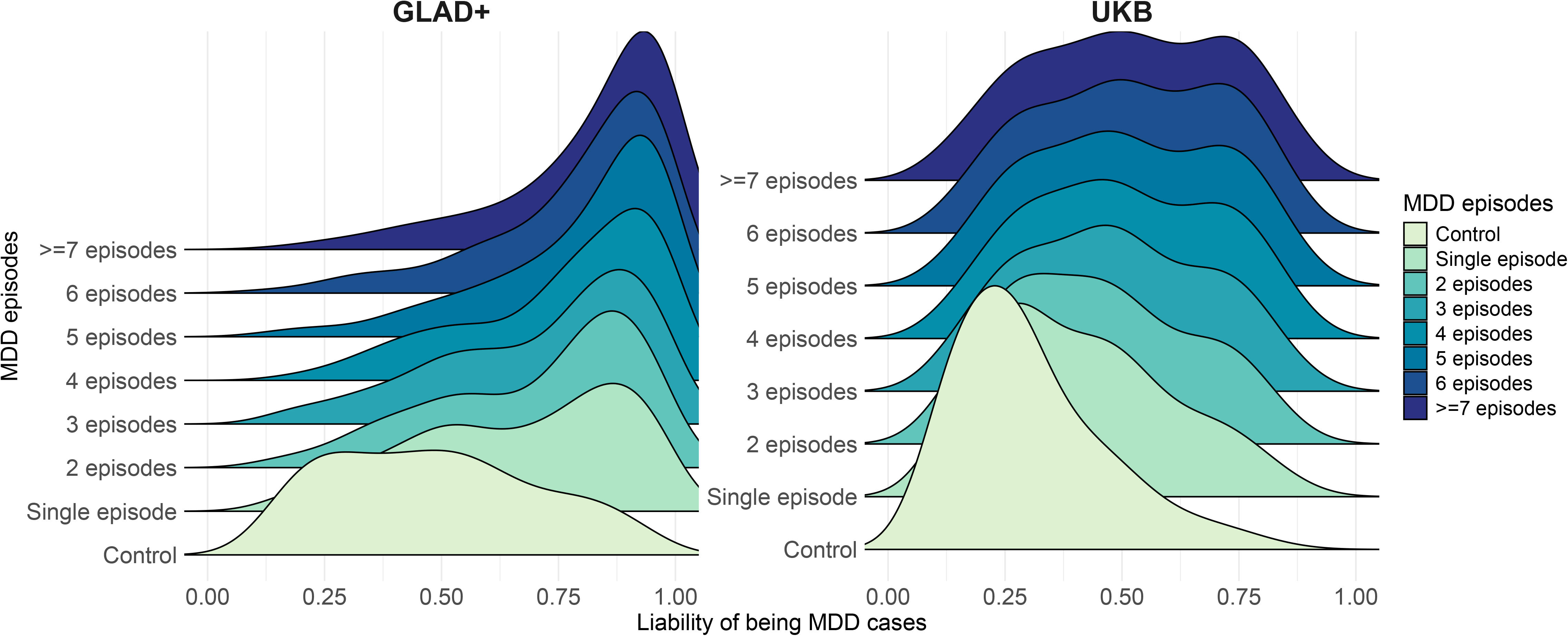
The distribution of the prediction score across frequency of MDD episodes in GLAD+ (n=12,876) and UKB (n=102,821) Alt text: Ridgeline density plots of the predicted MDD risk score for each category of lifetime episode count (from none to 13 or more) in GLAD+ (n=12,876) and UK Biobank (n=102,821). Predicted risk shifts upward with more episodes in both cohorts, but the distributions overlap substantially and the gradient is weaker in UK Biobank.

Models trained directly on the number of episodes explained 26.5% (GLAD+-trained) and 24.8% (UKB-trained) of the variance in GLAD+, and 12.1% (UKB-trained) and 10.1% (GLAD+-trained) in UKB (Table S20). The same predictors dominated (Figures S3-S4). For the number of episodes, the largest standardised coefficients were for childhood trauma (1.34 episodes per SD in GLAD+; 0.29 in UKB) and family history of depression (0.89; 0.37); for age of onset, childhood trauma (-1.71 and -2.03 years per SD), female sex (-1.75; -1.23), and family history of depression (-1.18; -1.53) predicted earlier onset.

Despite the attenuated cross-cohort R², observed episode counts increased consistently across prediction deciles in both cohorts (Figure S5), indicating preserved ranking; differences were mainly on the raw scale (Figure S6), consistent with calibration shifts. Resampling UKB cases to match the GLAD+ episode distribution improved the cross-cohort performance of the GLAD+-trained model for both MDD status (AUC 0.78, 95% CI 0.762-0.798 vs 0.74, 95% CI 0.737-0.743 in the full UKB sample) and the number of episodes (R²=14.5%, 95% CI 11.9-17.1% vs 10.1%, 95% CI 9.8-10.4%; Table S20), consistent with case-mix differences contributing to attenuation.

## Discussion

In two large UK cohorts spanning clinically enriched and population-based samples, integrating familial, genetic, and environmental risk factors gave consistent classification of lifetime MDD and of the number of depressive episodes. Family history and childhood trauma, the two risk factors a clinician can elicit in a single consultation, were stronger predictors than current PRSs, which contributed complementary but more modest information.

Family history outperformed mPRS in both cohorts (AUC 0.74 vs 0.66 in GLAD+; 0.65 vs 0.60 in UKB; Table S9), and the single-trait MDD PRS performed as well as the mPRS, so for depression the gain from combining PRSs across traits was small (Table S10). PRSs capture only common variants identified by GWASs, whereas family history reflects a broader set of familial risk factors: rare variants such as neurodevelopmental copy number variants and damaging coding variants(33, 34), non-additive genetic effects, gene-environment correlation, and shared family environment. Larger GWASs and sequencing studies will improve PRSs and may narrow the gap with SNP-based heritability(35), but a substantial difference between measured polygenic liability and overall familial risk remains. Family history of depression and the depression PRS were only weakly correlated (Pearson’s r = 0.10-0.15, Table S18), consistent with their being complementary rather than interchangeable markers of inherited susceptibility(10); combining them captures partially distinct components of inherited risk.

Predictors of diagnosis and of illness course were largely shared: childhood trauma and family history of depression were the strongest predictors of case status, number of episodes, and earlier onset in both cohorts (Figures S2-S4). The contribution of childhood trauma relative to family history was greatest for episode count in GLAD+, consistent with evidence linking trauma exposure to earlier onset and treatment-resistant depression(17). The diagnosis-trained model explained 27% of the variance in the number of episodes in GLAD+, equivalent to a correlation of 0.52, a very large effect by the benchmarks of psychological research(36), although estimates were lower in UKB. Prediction of treatment resistance was minimal, so current multivariable risk prediction cannot yet identify individuals at risk of poor treatment response.

Models trained in GLAD+ generalized to UKB, with attenuation that partly reflects differences in case mix; harmonizing the episode distribution partially restored performance (AUC 0.78 vs 0.74; Table S20). External validation also revealed reciprocal miscalibration, with UKB-trained models underestimating risk in GLAD+ and GLAD+-trained models overestimating it in UKB, in keeping with the difference in prevalence (69% vs 36%) and severity. Relative risk ranking is therefore transportable across cohorts, but accurate absolute risk estimation requires recalibration to the target population.

Clinically, these findings indicate that systematic assessment of family history and childhood trauma outperforms current PRSs for depression risk stratification; even without PRSs, the models retained strong performance (AUC 0.73-0.82, Table S11). Family history aggregates genetic and shared environmental risk, retrospectively reported childhood trauma is a powerful vulnerability marker, and both are readily obtained in routine practice. This supports a biopsychosocial view of risk in which inherited liability, early adversity, and demographic factors act together, and which is best captured by combining them rather than relying on any one. mPRS alone showed modest but above-chance discrimination (AUC 0.66 in GLAD+; 0.60 in UKB), which may become relevant where genomic data are held in medical records but family history is unknown or incomplete. The incremental gain from adding current PRSs was modest, however, and these results do not support routine clinical implementation of PRSs for depression at this stage, although their performance is likely to improve substantially(35). Because the models were developed and validated for lifetime MDD, their performance for incident MDD should be established in prospective cohorts before prognostic clinical application.

Threshold-based metrics add clinically interpretable information beyond AUC. In the clinically enriched GLAD+ sample, the high PPV suggests value for identifying likely cases but limited rule-out capacity; in the population-based UKB sample, the higher NPV suggests greater usefulness for identifying lower-risk individuals (Table S15). These contrasts primarily reflect the difference in prevalence (Table S16). In practice, the model would first be recalibrated to local prevalence through intercept adjustment(37), with the decision threshold set by the clinical goal and the relative cost of false-positive and false-negative results(38): for example, a high-NPV threshold for primary care triage, or a high-PPV threshold for case identification in specialist settings.

Several limitations should be considered. Analyses were restricted to participants of European genetic ancestry. Family history, childhood trauma, age at onset, and number of episodes were self-reported and subject to recall bias and misclassification. Family history was collected through an optional survey in GLAD+ (any relative) and in MHQ2 in UKB (first-degree relatives only), with differing participation biases, and missing family history was the main reason that 16% of otherwise eligible GLAD+ participants were excluded under complete-case analysis (Figure S1); information on the number, type, and degree of affected relatives could further improve prediction. GLAD+ and UKB differed in recruitment design, age structure, case prevalence, and severity; GLAD+ controls were older than cases, so age was not included as a predictor in either cohort, and the higher case proportion in GLAD+ may have influenced calibration. Case-control imbalance was addressed with class weights and case-mix differences with an exploratory resampling analysis. Genetic principal components were entered as penalized predictors; entering them unpenalized or regressing them out of the PRSs would adjust more robustly for population stratification. Finally, XGBoost used prespecified rather than tuned hyperparameters because the planned grid search did not complete, so its external performance may underestimate what a tuned non-linear model could achieve.

In summary, combining familial, genetic, and environmental risk factors gave robust and transportable prediction of MDD across clinically enriched and population-based cohorts. Family history and childhood trauma consistently outperformed PRSs, which added modest complementary information, and accurate absolute risk estimation required recalibration for prevalence and severity. These findings highlight both the promise and the current limitations of genetically informed prediction in depression: systematic assessment of family history and early adversity remains central to clinically meaningful risk stratification, independent of future genomic advances.

## Data and code availability

The GLAD+ Study data are available via a data access request application to the NIHR BioResource (https://bioresource.nihr.ac.uk/using-our-bioresource/academic-and-clinical-researchers/apply-for-bioresource-data/).

The code of the model build is available: https://github.com/RujiaWangEpi/MDD_prediction. The code for defining lifetime depression in MHQ2 in UKB is available: https://github.com/ColemanResearchGroup/MHQ2.

## Acknowledgements

We thank the GLAD+ Study, NIHR BioResource, and UK Biobank volunteers for their participation and gratefully acknowledge the contributions of NIHR BioResource centres, NHS Trusts, UK Biobank, and staff. We also thank the National Institute for Health & Care Research (NIHR), NHS Blood and Transplant, and Health Data Research UK as part of the Digital Innovation Hub Programme.

This study represents independent research funded by the NIHR Biomedical Research Centre at South London and Maudsley NHS Foundation Trust and King’s College London. Further information is available at https://www.maudsleybrc.nihr.ac.uk/facilities/bioresource/.

The views expressed are those of the authors and do not necessarily reflect those of the NHS, NIHR, HSC R&D Division, King’s College London, or the Department of Health and Social Care.

