## Supplement Materials for "Combining family history, polygenic scores, and childhood trauma to predict major depressive disorder"

**Supplementary materials**

**Supplementary Methods**

**Study samples**

**GLAD+ study**

The GLAD+ study includes two cohorts: the Genetic Links to Anxiety and Depression Study (GLAD) and the NIHR BioResource COVID-19 Psychiatric and Neurology Genetics study (COPING) in the UK. The GLAD Study ([www.gladstudy.org.uk](http://www.gladstudy.org.uk)) is an ongoing study that recruits participants who have experienced depression and/or anxiety online (via media campaigns). Participants provide demographic, environmental, and genetic data and consent to medical record linkage and potential recontact. Currently, the GLAD study has over 66,000 consented participants, with more than 50,000 having completed the online survey, and over 37,000 saliva samples collected. During the COVID-19 pandemic, the GLAD Study research team recontacted GLAD participants and healthy volunteers from other NIHR BioResource studies to conduct the COVID-19 Psychiatric and Neurology Genetics study (COPING). The goal was to estimate the impact of the COVID-19 pandemic on mental and neurological health. We recruited over 20,000 participants with common mental disorders and over 11,000 healthy volunteers, two-thirds of whom have been genotyped. All participants provided informed consent for the GLAD and/or COPING studies and the NIHR BioResource (<https://bioresource.nihr.ac.uk/>).

**UK Biobank**

The UK Biobank (UKB) is a prospective health study of over 500,000 individuals in the UK, aged 40-69 at recruitment (2006-2010). In 2016-17, participants were invited via email and postal newsletter to complete the first online Mental Health Questionnaire (MHQ1), resulting in responses from over 157,000 individuals by July 2017^1^. In 2022, the second Mental Health Questionnaire (MHQ2) was sent out to gather repeat data, new data, and data from participants who had not completed the first questionnaire. This led to responses from over 169,000 individuals in October-November 2022, including over 110,000 participants who had completed the earlier questionnaire (MHQ1 in 2016) and over 52,000 who had not. Consequently, more than 206,000 UK Biobank participants have data for either MHQ1 or MHQ2. Both MHQ1 and MHQ2 measured lifetime depression, anxiety disorders, and other psychiatric disorders. In MHQ2, self-reported family history of psychiatric disorders among first-degree relatives was collected. All participants provided informed consent for the UKB.

**Major depressive disorder (MDD)**

Lifetime major depressive disorder was assessed using the Adapted Composite International Diagnostic Interview-Short Form (CIDI-SF) in GLAD+ and UKB. According to the Diagnostic and Statistical Manual of Mental Disorders Fifth Edition (DSM-5) criteria, MDD cases were defined as individuals who have five or more of the following symptoms, with at least one of the core symptoms occurring daily or nearly every day for a duration of two weeks or more. The two core symptoms include depressed mood and loss of interest. The additional symptoms include sleep difficulties (insomnia or hypersomnia), weight changes, fatigue, diminished ability to think or concentrate, feelings of worthlessness and suicidal ideation.

In GLAD+, lifetime MDD was assessed in the COPING survey, which was sent to 47,127 GLAD and NIHR BioResource (NBR) participants. Of these, 20,191 met criteria for MDD and 11,064 were classified as controls. Controls were drawn from NBR only: GLAD participants not meeting MDD criteria (n = 2,072) were not used as controls, as the GLAD Study recruits individuals with depression and/or anxiety. We further excluded 2,717 NBR participants who did not meet MDD criteria but reported at least one depressive episode lasting two weeks or more, leaving 8,347 controls before genotype-based exclusions (**Figure S1**).

In UKB, we combined depression diagnoses across MHQ1 and MHQ2 using the same criteria. Participants meeting criteria for MDD in either MHQ1 (n=37,410) or MHQ2 (n=32,149) were defined as cases (n=54,670). Controls had not self-reported a diagnosis of depression, did not report either core symptom, and/or had a Patient Health Questionnaire-9 (PHQ-9) total score below 5, in both waves or in the only wave they completed (n=126,381). Controls who reported depressive episodes were excluded (n=38,719). The analytic sample was further restricted to participants of European genetic ancestry whose genotype data passed quality control (see “Genetic data in GLAD+ and UK Biobank”) and who had complete data on all predictors. Because family history was collected only in MHQ2, this restricted the sample to MHQ2 respondents and accounted for most exclusions. The final UKB sample comprised 40,667 MDD cases and 70,755 controls. The number excluded at each of these later steps could not be retrieved, because UKB individual-level data were inaccessible during revision. Participants of both studies were removed from GLAD+ rather than UKB (Figure S1), so this step did not affect the UKB sample.The CIDI-SF items in MHQ1 and MHQ2 are shown in **Table S1**.

**Table S1. CIDI-SF Items in MHQ1 and MHQ2 of the UKB**

| **MDD (CIDI-SF)** | **MHQ1** | **MHQ2** |
| --- | --- | --- |
| **2 core symptoms** |  |  |
| depressed mood | 20446 | 29011 |
| loss of interest | 20441 | 29012 |
| **additional symptoms** |  |  |
| sleep difficulties | 20532 | 29022 |
| weight changes | 20536 | 29021 |
| fatigue | 20449 | 29018 |
| diminished ability to think or concentrate | 20435 | 29026 |
| feelings of worthlessness | 20450 | 29027 |
| suicidal ideation | 20437 | 29029 |

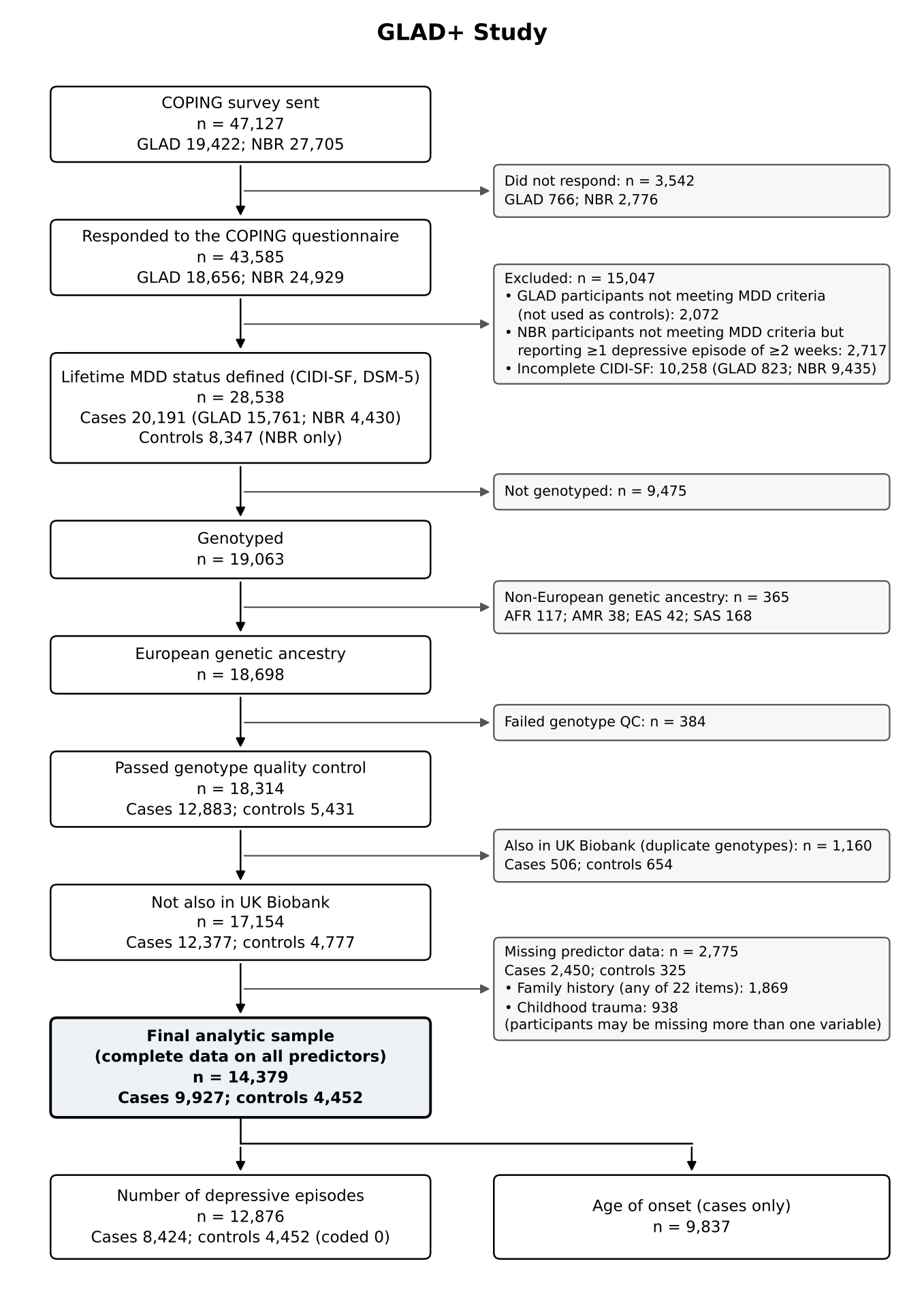

**Figure S1. Flow of participants in the GLAD+ Study**

Lifetime MDD was assessed with the CIDI-SF (DSM-5 criteria) in the COPING survey, which was sent to participants of the GLAD Study and the NIHR BioResource (NBR). Controls were NBR participants who did not meet criteria for lifetime MDD and reported no depressive episode lasting two weeks or more; GLAD participants not meeting MDD criteria were not used as controls, as the GLAD Study recruits individuals with depression and/or anxiety. Genetic ancestry was assigned by projection onto 1000 Genomes reference populations before genotype quality control (QC). Of 1,633 GLAD+ participants identified in UK Biobank through duplicate genotypes, 1,160 remained eligible at this step and were excluded. Participants were required to have complete data on all predictors, including all 22 family history items; participants may have been missing more than one variable. For number of depressive episodes, controls were coded as having no episodes. AFR, African; AMR, admixed American; CIDI-SF, Composite International Diagnostic Interview–Short Form; EAS, East Asian; EUR, European; MDD, major depressive disorder; NBR, NIHR BioResource; QC, quality control; SAS, South Asian.

**Number of MDD episodes**

The number of MDD episodes was assessed with the question: “Please estimate the number of times you have had periods of depression or low mood in your life lasting two or more weeks.” In GLAD+, answers were recorded in 13 categories (1 to 12, and 13 or more). UKB asked a similar question about periods of feeling depressed for at least two weeks, recorded as a count (field 20442 in MHQ1 and 29033 in MHQ2). For comparability, UKB counts were mapped onto the GLAD+ categories, with counts of 13 or more assigned to the “13 or more” category (n = 443). For participants who answered in both MHQ1 and MHQ2, we used the average of the MDD episodes.

**Age of MDD onset**

Age of MDD onset was determined by asking participants their age during their first two-week period of depression in both cohorts. In UKB, the index codes for the MDD age of onset are 20433 in MHQ1 and 29034 in MHQ2.

For the severity analyses, controls were coded as having no episodes and included in the analysis of the number of episodes, whereas age of onset was analysed in MDD cases only. Participants with missing responses were excluded, giving analytic samples of 12,876 in GLAD+ (8,424 cases and 4,452 controls) and 102,821 in UKB (32,066 cases and 70,755 controls) for the number of episodes, and 9,837 (GLAD+) and 38,395 (UKB) cases for age of onset.

**Treatment-resistant depression**

Treatment-resistant depression (TRD), according to the Maudsley Staging Method (MSM, Table S2), was assessed using a combination of the 4-item MSM and the depression severity score measured by the PHQ-9 in GLAD+. The PHQ-9 severity scoring categorizes depression as follows: 0-4 as none (0), 5-9 as mild (1), 10-14 as moderate (2), 15-19 as moderately severe (3), and 20-27 as severe (4). These scores can be recoded into a scale of 0-4. Participants were only shown the MSM if they had mild or more severe depression based on the PHQ-9 items, with scores ranging from 0 to 10. The MSM-TRD sum score combined the MSM score and PHQ-9 score, ranging from 0 to 14.

**Table S2. Maudsley Staging Method four-items**

| **CODE** | **Questions** | **Answers** |
| --- | --- | --- |
| MSM-1:  when low mood begin | How long ago did your current or most recent episode of depression or low mood begin? | 1= Less than 1 year ago  2= 1-2 years ago  3= More than 2 years ago |
| MSM-2:  taking antidepressants for 6 weeks or longer | During the current or most recent episode of depression or low mood, how many antidepressant medications have you taken for 6 weeks or longer? | 0= None  1= One to two  2= three to four  3= five to six  4= seven to ten  5= More than 10 |
| MSM-3:  add-on medication | If individuals don’t respond fully to antidepressants, doctors sometime prescribe “add-on” or “augmentation” medications in addition to the antidepressant (such as lithium, quetiapine or aripiprazole).During the current or most recent episode of depression or low mood, have you taken an add on medication for 6 weeks or longer? | 0= No  1= Yes |
| MSM-4:  ECT | Have you received electroconvulsive therapy (ECT) in this current or most recent episode of depression or low mood? (Please only answer yes if your course of ECT included 8 treatment sessions or more.) | 0= No  1= Yes |

**Family history (FH)**

In GLAD+, family history was assessed in the COPING survey by asking participants, “Have any of your family members ever been diagnosed with one or more of the following mental health disorders by a professional, even if they don’t have it currently?” The 22 response options were depression, depression during or after pregnancy, premenstrual dysphoric disorder (PMDD), bipolar disorder (BD), generalized anxiety disorder (GAD), social anxiety, specific phobia, agoraphobia, panic attacks, panic disorder (PD), post-traumatic stress disorder (PTSD), obsessive-compulsive disorder (OCD), body dysmorphic disorder (BDD), schizophrenia (SCZ), psychosis or psychotic illness, personality disorder, autism spectrum disorder (ASD), attention deficit/hyperactivity disorder (ADHD), anorexia nervosa (AN), atypical anorexia nervosa, bulimia nervosa (BN) and binge-eating disorder (BED) (**Table S3**). This question was answered by 32,776 of the 47,127 participants sent the COPING survey (GLAD 15,256; NBR 17,520). Each disorder was coded as a binary indicator (any affected family member: yes/no). For external validation, the GLAD+ items were mapped to the nine disorders assessed in UKB (**Table 1**).

In UKB, family history was assessed by asking participants, “Have any of your first degree blood relatives had any of the following conditions?” in MHQ2 (Index code 29001). The condition options available included depression, BD, anxiety disorder, SCZ, psychosis, personality disorder, ASD, ADHD, and eating disorder (ED) (Table 1). Participants who responded to the provided options for specified mental health disorders were considered to have a family history of corresponding mental health disorders. For the external dataset validation, family history of psychiatric disorders in GLAD+ were mapped the same as in UKB.

**Table S3. Characteristics of 22 family history predictors in GLAD+**

| **Family history**  (FH, yes, %) | **Total**  (n=14,379) | **MDD cases** (n=9,927) | **Controls**  (n=4,452) |
| --- | --- | --- | --- |
| depression | 7,116 (49.5) | 6,119 (61.6) | 997 (22.4) |
| depression during or after pregnancy | 1,440 (10.0) | 1,309 (13.2) | 131 (2.9) |
| premenstrual dysphoric disorder | 92 (0.64) | 88 (0.89) | 4 (0.09) |
| bipolar disorder | 1,219 (8.5) | 1,094 (11.0) | 125 (2.8) |
| generalized anxiety disorder | 4,139 (28.8) | 3,612 (36.4) | 527 (11.8) |
| social anxiety | 935 (6.5) | 868 (8.7) | 67 (1.5) |
| agoraphobia | 356 (2.5) | 333 (3.4) | 23 (0.52) |
| specific phobia | 321 (2.2) | 297 (3.0) | 24 (0.54) |
| panic attacks | 1,626 (11.3) | 1,441 (14.5) | 185 (4.2) |
| panic disorder | 608 (4.2) | 567 (5.7) | 41 (0.92) |
| PTSD | 696 (4.8) | 650 (6.5) | 46 (1.0) |
| SCZ | 596 (4.1) | 528 (5.3) | 68 (1.5) |
| ADHD | 647 (4.5) | 572 (5.8) | 75 (1.7) |
| ASD | 1,177 (8.2) | 1,021 (10.3) | 156 (3.5) |
| OCD | 644 (4.5) | 581 (5.9) | 63 (1.4) |
| BDD | 174 (1.2) | 164 (1.7) | 10 (0.22) |
| Psychosis | 584 (4.1) | 525 (5.3) | 59 (1.3) |
| Personality disorder | 475 (3.3) | 443 (4.5) | 32 (0.72) |
| Anorexia nervosa | 581 (4.0) | 498 (5.0) | 83 (1.9) |
| Atypical anorexia nervosa | 36 (0.25) | 33 (0.33) | 3 (0.07) |
| Bulimia nervosa | 301 (2.1) | 267 (2.7) | 34 (0.76) |
| Binge-eating disorder | 139 (0.97) | 130 (1.3) | 9 (0.20) |

Abbreviation: PTSD, post-traumatic stress disorder; OCD, obsessive-compulsive disorder; BDD, body dysmorphic disorder; SCZ, schizophrenia; ASD, autism spectrum disorder; ADHD, attention deficit / hyperactivity disorder.

**Genetic data in GLAD+ and UK Biobank**

**GLAD+**

All data from GLAD+ study were genotyped by ThermoFisher on the UK Biobank Axiom Array v1 and v2 across numerous genotyping batches. Genetic data for GLAD+ study were restricted to individuals from European ancestries (749,044 SNPs before quality control). Ancestry was determined using GenoPred ( <https://opain.github.io/GenoPred/index.html>) (GenoPredPipe, January 2024 version) , by projecting GLAD+ individuals on genomic principal components from the 1000 Genomes reference data, and assigning individuals a genetic ancestry if they lay < 3 SD from the mean of individuals from that ancestry superpopulation in 1000 Genomes. Quality control was conducted in PLINK v1.90b6.10 and v2.00a5.10, excluding variants with MAF < 0.01, call rate < 0.95, or which were deviant from Hardy-Weinberg equilibrium (p < 10^-10). Individuals were excluded if they had withdrawn from the study following genotyping, if they were a duplicate of a higher-quality sample (not including known identical twins), if they were known to be mislabelled, if their genotypic sex (males Fx > 0.8, females Fx < 0.5) did not match their sex assigned at birth, if they were outliers on genome-wide heterozygosity (absolute(Fhat) > 0.2), or if they had an excess of relatives (average pi-hat > 3 SD from the mean). Following quality control, 33,635 individuals and 484,182 variants were available for imputation. Imputation was carried out to TopMED Freeze 8, using the dedicated imputation server ( <https://imputation.biodatacatalyst.nhlbi.nih.gov/#!>). Following imputation, data was further restricted to data with MAF >= 0.01 and R2 >= 0.3, leaving 15,009,228 variants for analysis.

**UK Biobank**

UKB participants were genotyped on either the UK Biobank Axiom array (approximately 90% of participants) or the UK BiLEVE Axiom array^2^. Sample quality control was performed centrally by UK Biobank and included exclusions for outlying missingness or heterozygosity and for sex discordance^3^. UK Biobank provided imputed dosages for 487,422 participants based on 670,739 genotyped markers; imputation was performed with IMPUTE4 using the Haplotype Reference Consortium^4^, 1000 Genomes phase 3^5^ and UK10K^6^ reference panels. Imputed variants with a minor allele frequency < 0.01 or INFO score < 0.4 were excluded. Genetic ancestry was assigned with GenoPred (GenoPredPipe, January 2024 version; <https://opain.github.io/GenoPred/>)^7^ by projecting UKB participants onto genomic principal components derived from the 1000 Genomes reference data. Participants within 3 SD of the mean of a 1000 Genomes superpopulation were assigned to that ancestry, yielding 454,651 participants of European ancestry.

**MegaPRS**

In both cohorts, PRSs were generated for seven traits (MDD, anxiety, ASD, ADHD, bipolar disorder, PTSD and BMI) using GWAS summary statistics from participants of European ancestry that excluded UK Biobank samples; these PRSs were used in the main analyses. For a supplementary GLAD+-only analysis, PRSs were also generated for 15 further psychiatric disorders and related traits, some based on GWAS including UK Biobank samples (Table S4). PRSs were calculated with MegaPRS^8^ within the GenoPred pipeline^7^, using the BLD-LDAK heritability model, in which the variance explained by each SNP depends on its allele frequency, linkage disequilibrium and functional annotations. QC, model selection and standardisation are described below (GenoPred pipeline).

**GenoPred pipeline: summary statistic QC and harmonisation**

PRSs were generated with the GenoPred pipeline^7^ (GenoPredPipe, January 2024 version). Variants were matched between summary statistics, reference and target data by chromosome, base-pair position and alleles, using HapMap3 variants in the (1000 Genomes Phase 3) European reference.

*Summary statistic QC.* Variants were removed if they:

●	had an INFO score < 0.9 (where available);

●	had a minor allele frequency < 0.01;

●	had an allele frequency differing from the reference by > 0.2;

●	were strand-ambiguous (A/T or C/G) or duplicated;

●	had P-values outside the range 0-1 or a standard error of 0;

●	had a per-variant sample size more than 3 SD from the median (where available).

*Target data.* Imputed genotypes were converted to hard calls and restricted to the reference HapMap3 variants; reference variants missing in the target were imputed using the reference allele frequency.

*PRS calculation.* MegaPRS was run with LDAK v5.1 under the BLD-LDAK heritability model, which fits lasso, ridge, Bolt and BayesR models across 148 hyperparameter settings. The best model was selected by pseudo-validation using only the GWAS summary statistics, with the ancestry-matched reference as the LD reference. The MHC region was retained.

*Standardisation.* Scores were standardised using the mean and SD of the score in the ancestry-matched (European) reference sample, placing PRSs on the same scale in GLAD+ and UKB.

**Table S4. Sumstats for psychiatric disorders and related traits**

| **Disorders**  **/traits** | **Sumstats** | **PMID/ doi** | **N** |
| --- | --- | --- | --- |
| Depression * | MDD PGC3 without UK sample 2024 | 39814019 | Cases: 347,453;  Controls: 1,241,648 |
| Anxiety * | Anxiety PGC2 without UKB 2024 | <https://doi.org/10.1101/2024.07.03.24309466> | Cases: 94,930; Controls: 655,768 |
| Bipolar disorder * | Bipolar disorder without UKB sample 2021 | 34002096 | Cases: 41,917; Controls: 371,549; -UKB sample |
| Neuroticism | NEURO 2018 | 29500382 | 380,506 |
| Schizophrenia | SCZ 2022 | 35396580 | Cases: 53,386; Controls: 77,258 |
| ADHD * | PGC + iPSYCH 2023 | 36702997 | Cases: 38,691; Controls: 186,843 |
| ASD * | PGC + iPSYCH | 30804558 | Cases: 18,381; Controls: 27,969 |
| PTSD * | PTSD MVP 2021 | 33510476 | Cases: 36,301; Controls: 178,107 |
| Panic disorder | Panic 2019 | 31712720 | Cases: 2,147; Controls: 7,760 |
| AN | AN PGC2 | 31308545 | Cases: 16,992; Controls: 55,525 |
| OCD | PGC 2017 | 28761083 | Cases: 2,688;  Control: 7,037 |
| BMI * | GIANT 2015 | 25673413 | 322,154 |
| Educational attainment | EA 2022 | 35361970 | 3,037,499 |
| Alcohol dependence | Alcohol dependence 2017 | 30482948 | Cases: 11,569; Controls: 34,999 |
| Ever smoking | GSCAN-23andMe 2019 | 30643251 | 632,802 |
| Heavy smoking | UKB 2017 | 28166213 | 46,758 |
| Cannabis use | PGC 2020 | 33096046 | Cases: 17,068; Controls: 357,219 |
| Tiredness | UKB 2017 | 28322280 | 108,976 |
| Insomnia | Insomnia 2019 | 30804565 | 1,331,010 |
| Subjective well-being | SSGAC 2016 | 27089181 | 298,420 |
| Household income | UKB 2016 | 27818178 | 112,151 |
| Social deprivation | UKB 2016 | 27818178 | 112,151 |

*Note:* All discovery GWAS summary statistics were derived from participants of European ancestry. * PRSs generated in both GLAD+ and UKB and included in the main analyses; for these traits, summary statistics excluding UK or UKB samples were used. All other PRSs were generated in GLAD+ only.

**Childhood trauma (ChT)**

Reported childhood trauma was assessed using the Childhood Trauma Survey^9^, which includes five items measured on a Likert scale. Each item pertains to the frequency of a specific type of abuse (emotional, physical, sexual) or neglect (emotional, physical). Participants who reported experiencing at least one of these five types of childhood trauma were classified as individuals who had reported childhood trauma. Details of the five items assessing childhood trauma are shown in Table S5.

**Table S5. Items and definitions of childhood trauma**

| **Childhood trauma** | **Questions** | **Definition** | **MHQ1&MHQ2** |
| --- | --- | --- | --- |
| Emotional abuse | I felt that someone in my family hated me | If the answer is “Never true” or “Rarely true”, defined as no emotional abuse;    If the answer is “Sometimes true” or “Often true” or “Very often true”, it is defined as having emotional abuse. | 20487 & 29078 |
| Emotional neglect | When I was growing up, I felt loved | If the answer is “Sometimes true” or “Often true” or “Very often true”, defined as no emotional neglect;    If the answer is “Never true” or “Rarely true”, it is defined as having emotional neglect. | 20489 & 29076 |
| Physical abuse | People in my family hit me so hard that it left me with bruises or marks | If the answer is “Never true” or “Rarely true”, defined as no physical abuse;    If the answer is “Sometimes true” or “Often true” or “Very often true”, it is defined as having physical abuse. | 20488 & 29077 |
| Physical neglect | There was someone to take me to the doctor if I needed it | If the answer is “Sometimes true” or “Often true” or “Very often true”, defined as no physical neglect;    If the answer is “Never true” or “Rarely true”, it is defined as having physical neglect. | 20491 & 29080 |
| Sexual abuse | Someone molested me (sexually) | If the answer is “Never true”, defined as no sexual abuse;    If the answer is “Rarely true” or “Sometimes true” or “Often true” or “Very often true”, it is defined as having sexual abuse. | 20490 & 29079 |

**Prediction model**

**Software**

Elastic net models were fitted in R 4.3.0 using caret (v7.0-1; method = “glmnet”) and glmnet (v4.1-8), with parallel processing via doMC (v1.3.8). AUCs were computed with pROC (v1.18.5) and Brier scores with verification (v1.44). XGBoost models and DeLong tests were run in R 4.4.0 using xgboost (v3.1.3.1) and pROC (v1.19.0.1). All versions are listed in Table S21 and random seeds in Table S6.

**Missing data**

Predictors missing in ≥10% of participants were to be excluded from modelling; no predictor met this threshold. Participants with missing data on any remaining predictor (including all 22 family-history items in GLAD+) or on the outcome were then removed. Elastic net and XGBoost models were fitted to the same complete-case data (Figure S1).

**Class weighting for imbalanced sample**

Case-control imbalance was addressed with class weights, with each case weighted by the ratio of controls to cases and each control weighted 1. For the elastic net, weights were computed within each outer training fold and applied only to the training loss; held-out folds were evaluated without weights. Because outer folds were randomly assigned, the ratio varied slightly between folds around the full-sample values: 9,927 cases and 4,452 controls in GLAD+ (case weight ≈ 0.45, equivalent to weighting controls 2.23 relative to cases) and 40,667 cases and 70,755 controls in UKB (case weight ≈ 1.74). For XGBoost, scale_pos_weight was set to the full-sample ratio of controls to cases (GLAD+ 0.45; UKB 1.74); because XGBoost folds were stratified by case status, this equals the ratio in each training fold (Table S6).

**XGBoost**

To assess potential non-linear relationships among predictors, we implemented gradient-boosted decision trees using XGBoost (v3.1.3.1) in R 4.4.0 as a secondary modelling approach. XGBoost models used the same predictor sets as the primary elastic net analyses. A grid search over 18 hyperparameter combinations (learning rate 0.03, 0.05 or 0.1; maximum depth 3, 4 or 6; minimum child weight 1 or 5) with cross-validated early stopping was planned, but it did not return a valid result in any analysis, and the script used its prespecified fallback values: learning rate 0.05, maximum depth 4, minimum child weight 1, 200 boosting rounds, row and column subsampling 0.8, and default regularisation (gamma 0, L1 0, L2 1; Table S6). Class imbalance was handled by setting scale_pos_weight to the full-sample ratio of controls to cases; because folds were stratified by case status, this equals the ratio in each training fold. Within each cohort, performance was evaluated using 5-fold cross-validation stratified by outcome (random seed 2026). Models were trained on four folds and used to predict the held-out fold, and discrimination was summarised as the mean AUC (±SD) across folds. Calibration intercept and slope were estimated from out-of-fold predicted probabilities using a logistic recalibration model, alongside the Brier score. For external validation, models were trained on the full development cohort and applied without re-estimation to the other cohort.

**Table S6. Hyperparameter specification and selected values for each learner**

|  | **Elastic net** | **XGBoost** |
| --- | --- | --- |
| Software | R 4.3.0; caret 7.0-1 (method = “glmnet”); glmnet 4.1-8 | R 4.4.0; xgboost 3.1.3.1 |
| Hyperparameters | Tuned: mixing parameter (α) and penalty strength (λ) | Not tuned: the specified grid search did not complete, so prespecified default values were used |
| Search space | α ∈ {0.10, 0.55, 1.00}; three λ values from the glmnet regularization path (caret default grid; 9 combinations) | Planned: learning rate {0.03, 0.05, 0.1} × maximum depth {3, 4, 6} × minimum child weight {1, 5} (18 combinations); not applied |
| Fixed parameters | — | booster = gbtree; objective = binary:logistic; eta = 0.05; max_depth = 4; min_child_weight = 1; boosting rounds = 200; subsample = 0.8; colsample_bytree = 0.8; gamma = 0; L1 (alpha) = 0; L2 (lambda) = 1 |
| Search method | Grid search | Not applicable |
| Tuning criterion | Classification accuracy (binary outcomes); RMSE (continuous outcomes) | Not applicable |
| Evaluation criterion | AUC (binary); R² (continuous) | AUC |
| Cross-validation | Nested: 10-fold inner CV for tuning (stratified by outcome) within 5-fold outer CV for evaluation | 5-fold CV stratified by outcome, for evaluation only |
| Class imbalance | Case weight = controls/cases, computed within each training fold | scale_pos_weight = controls/cases in the full sample (GLAD+: 0.45; UKB: 1.74); equals the training-fold ratio because folds were stratified |
| Random seed | 1 | 2026 |
| Selected values, GLAD+ (MDD) | α = 0.1-1.0 across folds; λ = 0.0003-0.033; 26-28 of 28 predictors retained | Fallback values in all analyses (no tuning AUC recorded; 200 rounds) |
| Selected values, UKB (MDD) | α = 1.0 in all folds; λ = 0.029 (range 0.0285-0.0292); 22-25 of 28 predictors retained | Fallback values in all analyses (no tuning AUC recorded; 200 rounds) |

*Note.* CV, cross-validation; RMSE, root mean squared error; AUC, area under the receiver operating characteristic curve. Values are for the harmonized predictor set (9 family history indicators, 7 PRSs, childhood trauma, sex and 10 genetic principal components). Selected values by outer fold are shown in Table S6a.

**Table S6a. Elastic net hyperparameters and retained predictors by outer fold (MDD, episodes and age of onset)**

| **Phenotype** | **Cohort** | **Outer fold** | **Training N** | **α** | **λ** | **Non-zero predictors** |
| --- | --- | --- | --- | --- | --- | --- |
| **MDD** | GLAD+ | 1 | 11,503 | 0.55 | 0.0033 | 26 / 28 |
|  |  | 2 | 11,503 | 0.1 | 0.0332 | 28 / 28 |
|  |  | 3 | 11,503 | 0.55 | 0.0034 | 27 / 28 |
|  |  | 4 | 11,503 | 0.1 | 0.00033 | 27 / 28 |
|  |  | 5 | 11,504 | 1.0 | 0.0034 | 26 / 28 |
|  | UKB | 1 | 89,138 | 1.0 | 0.0292 | 25 / 28 |
|  |  | 2 | 89,138 | 1.0 | 0.0285 | 23 / 28 |
|  |  | 3 | 89,138 | 1.0 | 0.0289 | 22 / 28 |
|  |  | 4 | 89,137 | 1.0 | 0.0288 | 23 / 28 |
|  |  | 5 | 89,137 | 1.0 | 0.0286 | 25 / 28 |
| **Episodes** | GLAD+ | 1 | 10,302 | 0.55 | 0.0371 | 27 / 28 |
|  |  | 2 | 10,301 | 0.55 | 0.0369 | 26 / 28 |
|  |  | 3 | 10,300 | 0.55 | 0.0376 | 28 / 28 |
|  |  | 4 | 10,300 | 0.55 | 0.0369 | 26 / 28 |
|  |  | 5 | 10,301 | 1.00 | 0.0370 | 27 / 28 |
|  | UKB | 1 | 82,257 | 0.10 | 0.0097 | 28 / 28 |
|  |  | 2 | 82,257 | 0.55 | 0.0097 | 27 / 28 |
|  |  | 3 | 82,257 | 0.55 | 0.0098 | 25 / 28 |
|  |  | 4 | 82,257 | 0.10 | 0.0098 | 27 / 28 |
|  |  | 5 | 82,256 | 0.10 | 0.0096 | 27 / 28 |
| **Age of onset** | GLAD+ | 1 | 7,869 | 0.10 | 0.450 | 28 / 28 |
|  |  | 2 | 7,871 | 1.00 | 0.042 | 28 / 28 |
|  |  | 3 | 7,870 | 0.10 | 0.402 | 27 / 28 |
|  |  | 4 | 7,869 | 0.10 | 0.433 | 28 / 28 |
|  |  | 5 | 7,869 | 0.10 | 0.420 | 28 / 28 |
|  | UKB | 1 | 30,716 | 0.55 | 0.0524 | 28 / 28 |
|  |  | 2 | 30,716 | 0.55 | 0.0520 | 28 / 28 |
|  |  | 3 | 30,717 | 0.55 | 0.0523 | 27 / 28 |
|  |  | 4 | 30,716 | 0.55 | 0.0535 | 27 / 28 |
|  |  | 5 | 30,715 | 0.55 | 0.0516 | 28 / 28 |

Note. Non-zero predictors: number of the 28 predictors with a non-zero coefficient in each outer-fold model. For MDD in GLAD+, 23 of the 28 predictors were retained in all five folds: childhood trauma, sex, all seven PRSs, eight of the nine family history indicators (family history of ADHD was retained in four folds) and six of the ten genetic principal components; the remaining four principal components were retained in three or four folds (Table S7). For MDD in UKB, 21 of the 28 predictors were retained in all five folds, and nine of the ten principal components were retained in at least one fold. For the number of episodes and age of onset, 23-27 predictors were retained in all five folds in both cohorts.

**Supplementary Results**

**Table S7. Coefficients of the elastic net model (all predictors) refit on the full development dataset, and their range across the five outer-fold models in GLAD+**

| **Group** | **Predictor** | **Full refit (100%)** | **Outer folds (80%)** | | **Selected in outer folds** |
| --- | --- | --- | --- | --- | --- |
|  |  | **Beta** | **Min Beta** | **Max Beta** |  |
| **mFH** | Depression | 1.146 | 1.126 | 1.233 | 5/5 |
|  | Anxiety | 0.658 | 0.635 | 0.704 | 5/5 |
|  | Mania | 0.420 | 0.383 | 0.518 | 5/5 |
|  | ADHD | 0.050 | 0.000 | 0.162 | 4/5 |
|  | Autism | 0.328 | 0.293 | 0.412 | 5/5 |
|  | Eating disorder | 0.384 | 0.332 | 0.499 | 5/5 |
|  | Personality disorder | 0.391 | 0.292 | 0.574 | 5/5 |
|  | Psychosis | 0.148 | 0.093 | 0.268 | 5/5 |
|  | Schizophrenia | 0.265 | 0.227 | 0.378 | 5/5 |
| **mPRS** | Depression | 0.328 | 0.311 | 0.358 | 5/5 |
|  | Anxiety | 0.121 | 0.107 | 0.144 | 5/5 |
|  | Autism | 0.048 | 0.047 | 0.073 | 5/5 |
|  | PTSD | 0.036 | 0.010 | 0.052 | 5/5 |
|  | Bipolar | 0.034 | 0.028 | 0.056 | 5/5 |
|  | ADHD | 0 (not selected) | -0.033 | 0.012 | 5/5 |
|  | Body mass index | 0.019 | 0.018 | 0.026 | 5/5 |
| **ChT** | Childhood trauma | 1.348 | 1.336 | 1.400 | 5/5 |
| **Background** | Sex | 1.100 | 1.099 | 1.144 | 5/5 |
|  | PC1 | 0.091 | 0.088 | 0.122 | 5/5 |
|  | PC2 | -0.027 | -0.060 | 0.000 | 3/5 |
|  | PC3 | 0 (not selected) | -0.023 | 0.013 | 4/5 |
|  | PC4 | 0.079 | 0.077 | 0.099 | 5/5 |
|  | PC5 | 0.028 | 0.020 | 0.043 | 5/5 |
|  | PC6 | 0 (not selected) | -0.017 | 0.010 | 4/5 |
|  | PC7 | 0 (not selected) | -0.016 | 0.019 | 5/5 |
|  | PC8 | 0.005 | 0.004 | 0.032 | 5/5 |
|  | PC9 | 0 (not selected) | -0.010 | 0.012 | 4/5 |
|  | PC10 | 0.072 | 0.070 | 0.098 | 5/5 |

Note: Beta, elastic net log-odds coefficient per unit of the predictor as entered (polygenic scores and principal components standardized; family history, childhood trauma and sex coded as in the main analysis). Full refit: model refit on the full development dataset, with α and λ selected by 10-fold cross-validation (the same procedure as the inner loop of the nested cross-validation). Outer folds: models trained on 80% of the development data within each outer fold of the nested cross-validation. Min/Max: range of the five fold-specific coefficients. 0 (not selected): coefficient shrunk to exactly zero by elastic net regularization. Selected in outer folds: number of outer-fold models with a non-zero coefficient. Models were trained with class weights to account for case-control imbalance; the intercept therefore does not reflect the observed prevalence, and predicted probabilities are not calibrated to population prevalence.

**Table S7a. Agreement between each outer-fold model and the full refit (28 predictors) in GLAD+**

| **Outer fold** | **Pearson r** | **Lin's CCC** | **Refit Beta within fold range** |
| --- | --- | --- | --- |
| Fold 1 | 0.998 | 0.998 | 28/28 |
| Fold 2 | 0.993 | 0.990 |  |
| Fold 3 | 0.996 | 0.996 |  |
| Fold 4 | 0.993 | 0.992 |  |
| Fold 5 | 0.994 | 0.993 |  |

CCC, concordance correlation coefficient (Lin), which measures absolute agreement with the identity line rather than linear association only. Refit β within fold range: number of the 28 predictors whose full-refit coefficient lies within the range of the five outer-fold coefficients.

**Table S8. Observed-scale and liability-scale R² for MDD status prediction across assumed population prevalences (K) in GLAD+ and UK Biobank**

| Dataset | Cohort | Case proportion | Observed R² (%)  (95% CI) | Liability R² (%) (95% CI) | | | | N |
| --- | --- | --- | --- | --- | --- | --- | --- | --- |
|  |  |  |  | K = 5% | K = 10% | K = 15% | K = 20% |  |
| Training | GLAD+ | 0.69 | 29.1  (27.9-30.4) | **32.2**  **(30.7-33.7)** | 37.9  (36.2-39.6) | 41.6  (39.7-43.3) | 44.2  (42.3-46.1) | 14,379 |
| Testing | UKB | 0.36 | 17.0  (16.5-17.5) | 17.3  (16.8-17.9) | 20.9  (20.2-21.5) | **23.2**  **(22.5-23.9)** | 24.9  (24.1-25.6) | 111,422 |
| Testing | UKB-  resampled | 0.65 | 20.5  (17.4-23.6) | 21.0  (17.5-24.5) | 25.1  (21.1-29.2) | **27.9**  **(23.5-32.3)** | 29.9  (25.3-34.6) | 2,442 |
| Training | UKB | 0.36 | 16.9  (16.4-17.4) | 17.2  (16.7-17.8) | 20.8  (20.1-21.4) | **23.1**  **(22.4-23.8)** | 24.8  (24.0-25.5) | 111,422 |
| Testing | GLAD+ | 0.69 | 27.2  (25.8-28.5) | **29.8**  **(28.2-31.4)** | 35.2  (33.4-37.0) | 38.7  (36.8-40.6) | 41.3  (39.3-43.3) | 14,379 |

Observed R² was back-calculated from the liability-scale R² reported in the original analysis (K = 5% for GLAD+; K = 15% for UK Biobank and resampled UK Biobank) and the case proportion of each validation sample, and then re-expressed on the liability scale at K = 5%, 10%, 15% and 20% (Lee et al., Genet Epidemiol 2012). Confidence intervals were transformed in the same way. Bold: values reported in the original analysis. Case proportion: GLAD+ 9,927/14,379; UK Biobank 40,667/111,422; resampled UK Biobank 1,597/2,442. Observed-scale R² do not depend on the assumed prevalence. Training: pooled out-of-fold predictions from nested cross-validation within the cohort. Testing: model trained in the other cohort and applied unchanged (UK Biobank and resampled UK Biobank: GLAD+-trained model; GLAD+: UK Biobank-trained model).

**Table S9. Pairwise comparisons of ROC-AUCs across predictor groups in GLAD+ and UK Biobank**

| **Cohort** | **Model1** | **Model2** | **AUC1** | **AUC2** | **ΔAUC** | **P value** | **FDR-**  **adjusted P-value** | **Bonferroni- adjusted P-value** |
| --- | --- | --- | --- | --- | --- | --- | --- | --- |
| **GLAD+** | FH | PRS | 0.733 | 0.655 | -0.078 | 2.46E-35 | 4.92E-35 | 2.46E-34 |
|  | FH | CT | 0.733 | 0.667 | -0.066 | 1.88E-26 | 3.13E-26 | 1.88E-25 |
|  | FH | Demo | 0.733 | 0.671 | -0.062 | 1.45E-22 | 2.07E-22 | 1.45E-21 |
|  | FH | All | 0.733 | 0.833 | 0.100 | 4.65E-180 | 1.16E-179 | 4.65E-179 |
|  | PRS | CT | 0.655 | 0.667 | 0.012 | 0.0683 | 0.0758 | 0.6826 |
|  | PRS | Demo | 0.655 | 0.671 | 0.016 | 0.0211 | 0.0263 | 0.2106 |
|  | PRS | All | 0.655 | 0.833 | 0.178 | 3.09E-290 | 1.55E-289 | 3.09E-289 |
|  | CT | Demo | 0.667 | 0.671 | 0.004 | 0.5197 | 0.5197 | 1 |
|  | CT | All | 0.667 | 0.833 | 0.166 | 1.24E-296 | 1.24E-295 | 1.24E-295 |
|  | Demo | All | 0.671 | 0.833 | 0.162 | 3.00E-243 | 9.99E-243 | 3.00E-242 |
| **UKB** | FH | PRS | 0.647 | 0.599 | -0.048 | 1.09E-89 | 2.18E-89 | 1.09E-88 |
|  | FH | CT | 0.647 | 0.609 | -0.038 | 3.18E-61 | 5.30E-61 | 3.18E-60 |
|  | FH | Demo | 0.647 | 0.620 | -0.028 | 3.93E-31 | 5.62E-31 | 3.93E-30 |
|  | FH | All | 0.647 | 0.738 | 0.091 | <2.2E-308 | <2.2E-308 | <2.2E-308 |
|  | PRS | CT | 0.599 | 0.609 | 0.010 | 1.80E-5 | 1.80E-5 | 1.80E-4 |
|  | PRS | Demo | 0.599 | 0.620 | 0.021 | 1.90E-17 | 2.37E-17 | 1.90E-16 |
|  | PRS | All | 0.599 | 0.738 | 0.139 | <2.2E-308 | <2.2E-308 | <2.2E-308 |
|  | CT | Demo | 0.609 | 0.620 | 0.011 | 1.42E-5 | 1.58E-5 | 1.42E-4 |
|  | CT | All | 0.609 | 0.738 | 0.129 | <2.2E-308 | <2.2E-308 | <2.2E-308 |
|  | Demo | All | 0.620 | 0.738 | 0.118 | <2.2E-308 | <2.2E-308 | <2.2E-308 |

Note: ΔAUC = AUC2 − AUC1. P-values from DeLong’s test for paired ROC curves (pROC v1.19.0.1), based on pooled out-of-fold predictions from identical outer folds. As DeLong’s test may be miscalibrated for nested models (Demler et al., 2012), comparisons with the full model (All) should be interpreted alongside ΔAUC. <2.2e-308: below the smallest value R can represent. FH, family history; PRS, polygenic risk scores; CT, childhood trauma; Demo, background factors (sex and ten genetic principal components); All, full model.

**Table S10. Predictive performance of single-trait MDD PRS and multi-trait PRS for MDD (nested elastic net)**

| **PRS predictors** | **GLAD+** | | **UKB** | |
| --- | --- | --- | --- | --- |
|  | **AUC** | **R²** | **AUC** | **R²** |
| MDD PRS (single trait) | 0.65 | 5.5% | 0.59 | 3.3% |
| mPRS (7 traits) | 0.66 | 6.2% | 0.60 | 3.6% |
| Difference (mPRS-MDD PRS) | 0.01 | 0.7% | 0.01 | 0.3% |

**Table S11. Leave out mPRS models in GLAD+ and UK Biobank**

| **Cohort** | **Dataset** | **Predictors** | **Liability R^2^** | **Liability R^2^ deduction** | **AUC** | **Proportion of AUC deduction** |
| --- | --- | --- | --- | --- | --- | --- |
| **GLAD+** | Training | All predictors | 32.2% |  | 0.83 |  |
|  | Training | Leave-out mPRS | 26.6% | 17.4% | 0.82 | 1.8% |
|  | Testing | Leave-out mPRS | 29.1% | 2.3% | 0.81 | 1.2% |
| **UKB** | Training | All predictors | 23.1% |  | 0.74 |  |
|  | Training | Leave-out mPRS | 20.8% | 10.0% | 0.73 | 1.4% |
|  | Testing | Leave-out mPRS | 20.5% | 11.6% | 0.72 | 2.7% |

The testing dataset represents external dataset testing. For GLAD+ testing, the model was trained on the UKB and tested on the GLAD+ study, and vice versa. Deduction = (full model − leave-out model) / full model, compared with the full model in the same training or testing setting (testing references: GLAD+ R² 29.8%, AUC 0.82; UKB R² 23.2%, AUC 0.74).

**Table S12. Incremental discrimination of each predictor domain over background factors for MDD in GLAD+ and UK Biobank**

| **Predictors in the model** | **GLAD+** | | **UKB** | |
| --- | --- | --- | --- | --- |
|  | **AUC** | **𝛥AUC** | **AUC** | **𝛥AUC** |
| Background (sex +10 PCs) | 0.67 | reference | 0.62 | reference |
| Background + mFH | 0.78 | 0.11 | – | – |
| Background + mPRS | 0.73 | 0.06 | 0.66 | 0.04 |
| Background + ChT | 0.75 | 0.08 | – | – |
| Full model (all predictors) | 0.83 | 0.16 | 0.74 | 0.12 |

*Note.* AUC from nested elastic net cross-validation. ΔAUC is relative to background factors in the same cohort. UK Biobank results for background factors alone, background + mPRS and the full model come from analyses completed before UK Biobank data access ended. Models combining background factors with mFH or ChT were added during revision and could not be fitted in UK Biobank, because UK Biobank data could not be accessed during revision (–). mFH = family history of multiple psychiatric disorders; ChT = childhood trauma; mPRS = multi-trait polygenic risk scores; PCs = genetic principal components.

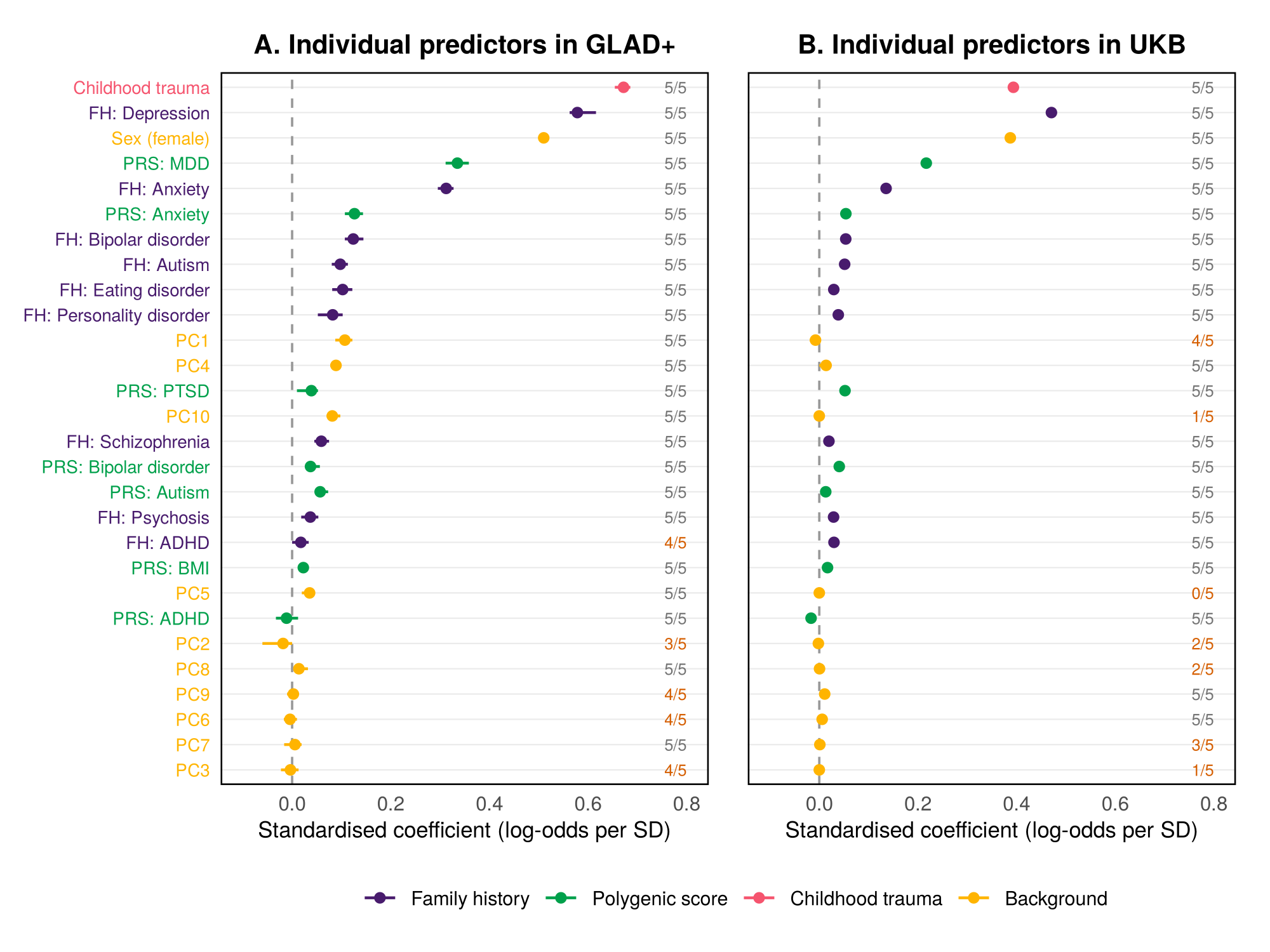

**Figure S2. Contribution of individual predictors to the elastic net model for MDD in GLAD+ and UK Biobank**

Standardised coefficients from the elastic net model including all predictors in GLAD+ (A) and UK Biobank (B). Points show the standardised coefficient (coefficient × predictor SD; change in log-odds of MDD per SD of the predictor), averaged across the five outer-fold models of the nested cross-validation; bars show the range across folds. Labels on the right give the number of outer folds in which the predictor had a non-zero coefficient (orange: fewer than five). Predictors are ordered by their mean absolute standardised coefficient across the two cohorts. Standard deviations of binary predictors were calculated as √[p(1−p)] from their prevalence (Table 1); polygenic scores and genetic principal components were standardised (SD = 1). Coefficients reflect each predictor’s contribution conditional on all other predictors in the penalised model and are shrunk towards zero. FH, family history, including family history of 9 psychiatric disorders in GLAD+ and UKB. PRS, polygenic risk scores, including 7 PRSs of psychiatric disorders and BMI based without UK/UKB GWAS sumstats in GLAD+ and UKB. Background, sociodemographic factors, including sex and 10 principal components.

**Table S13. Prediction of MDD status in GLAD+ using family history and polygenic risk scores for all 22 available traits compared with the traits mapped across GLAD+ and UK Biobank**

| **Predictors** | **Full predictors**  **(FH and PRS, 22 traits)** | | | **Mapped predictors**  **(FH for 9 traits and PRS for 7 traits)** | | | **𝛥AUC** | **P-value** |
| --- | --- | --- | --- | --- | --- | --- | --- | --- |
|  | **Beta (SE)** | **R²** | **AUC** | **Beta (SE)** | **R²** | **AUC** |  |  |
| mFH | 0.40 (0.008) | 16.6% | 0.740 | 0.39 (0.008) | 16.1% | 0.733 | 0.007 | 8.95E-11 |
| mPRS | 0.27 (0.008) | 7.3% | 0.666 | 0.25 (0.008) | 6.6% | 0.655 | 0.011 | 1.02E-07 |
| ChT | 0.32 (0.008) | 10.6% | 0.667 | 0.32 (0.008) | 10.6% | 0.667 | – | – |
| Background  (sex + 10 PCs) | 0.31 (0.008) | 9.8% | 0.672 | 0.31 (0.008) | 9.8% | 0.672 | – | – |
| All predictors | 0.55 (0.007) | 32.9% | 0.836 | 0.54 (0.007) | 32.2% | 0.833 | 0.003 | 5.92E-05 |

mFH, multi-trait family history score; mPRS, multi-trait polygenic risk score; ChT, childhood trauma; PCs, genetic principal components. 22 traits: family history and PRSs for all 22 traits available in GLAD+. Mapped traits: family history and PRSs for the 7 traits available in both GLAD+ and UK Biobank, as used in the main analysis. Models with 22 traits could not be validated externally because these traits were not available in UK Biobank. ChT and background factors are identical in both sets. Each row shows a logistic regression of MDD status on the predicted score from the indicated predictor set (N = 14,379; 9,927 cases and 4,452 controls). “All predictors” includes mFH, mPRS, ChT, sex and 10 PCs modelled jointly. Beta (SE): log-odds of MDD per standard deviation increase in the predicted score. R²: liability-scale R² assuming a population prevalence of 5% (Lee et al., Genet Epidemiol 2012). AUC: area under the receiver operating characteristic curve.

**Table S14. Calibration for MDD prediction in GLAD+ and UK Biobank**

| **MDD prediction** | | **Model** | **Brier score** | **Calibration slope** | **Calibration intercept** |
| --- | --- | --- | --- | --- | --- |
| **Train** | **Test** |  |  |  |  |
| GLAD+ | GLAD+ | Elastic net | 0.169 | 0.895 | 0.188 |
|  |  | XGBoost | 0.167 | 0.982 | 0.739 |
| UKB | UKB | Elastic net | 0.208 | 1.286 | -0.255 |
|  |  | XGBoost | 0.204 | 1.015 | -0.546 |
| GLAD+ | UKB | Elastic net | 0.197 | 0.793 | 0.038 |
|  |  | XGBoost | 0.309 | 0.846 | -1.446 |
| GLAD+ | UKB resample | Elastic net | 0.205 | 0.797 | 0.252 |
|  |  | XGBoost | 0.196 | 0.959 | -0.451 |
| UKB | GLAD+ | Elastic net | 0.174 | 1.399 | -0.114 |
|  |  | XGBoost | 0.166 | 1.401 | -0.703 |

Calibration performance of Elastic Net and XGBoost models for major depressive disorder prediction. Internal calibration was assessed within cohorts, while external calibration evaluated model transportability between GLAD+ and UK Biobank. An additional resampled UKB analysis was performed to account for differences in outcome prevalence. Brier score summarises overall accuracy; the calibration slope (ideal = 1) indicates whether predictions are too extreme (< 1) or too moderate (> 1), and the calibration intercept (ideal = 0) indicates systematic over- or underestimation. The resampled UKB sample was matched to the GLAD+ distribution of episodes.

**Table S15. PPV, NPV, sensitivity and specificity for prediction models in external validation GLAD+ and UK Biobank**

| **Cohort** | **Prediction score cut-off** | **0.3** | **0.4** | **0.5** | **0.6** | **0.7** |
| --- | --- | --- | --- | --- | --- | --- |
| **GLAD+ external validation** | PPV | 0.73 | 0.78 | **0.86** | 0.90 | 0.95 |
|  | NPV | 0.82 | 0.72 | **0.58** | 0.51 | 0.41 |
|  | Sensitivity | 0.98 | 0.93 | **0.77** | 0.64 | 0.38 |
|  | Specificity | 0.17 | 0.42 | **0.72** | 0.84 | 0.96 |
| **UKB external validation** | PPV | 0.49 | 0.55 | **0.60** | 0.65 | 0.71 |
|  | NPV | 0.81 | 0.78 | **0.76** | 0.73 | 0.71 |
|  | Sensitivity | 0.79 | 0.66 | **0.56** | 0.46 | 0.33 |
|  | Specificity | 0.53 | 0.69 | **0.79** | 0.86 | 0.92 |
| **UKB-**  **resample external validation** | PPV | 0.77 | 0.81 | **0.85** | 0.87 | 0.90 |
|  | NPV | 0.62 | 0.57 | **0.53** | 0.49 | 0.45 |
|  | Sensitivity | 0.83 | 0.73 | **0.63** | 0.54 | 0.40 |
|  | Specificity | 0.53 | 0.68 | **0.78** | 0.85 | 0.91 |

Note: GLAD+ results represent the model trained on UKB and tested for external validation in GLAD+. UKB and UKB-resample results represent the model trained on GLAD+ and tested for external validation in UKB and UKB resampled datasets.

UKB resample MDD prevalence 0.65

UKB MDD prevalence: 0.36

GLAD+ MDD prevalence: 0.69

**Table S16. Expected PPV and NPV by prevalence at the 0.5 threshold, from the observed sensitivity (Se) and specificity (Sp).**

| **MDD prevalence** | **Model validated in GLAD+**  **(Se 0.77, Sp 0.72): PPV/NPV** | **Model validated in UKB**  **(Se 0.56, Sp 0.79): PPV/NPV** |
| --- | --- | --- |
| 10% | 0.23 / 0.97 | 0.23 / 0.94 |
| 20% | 0.41 / 0.93 | 0.40 / 0.88 |
| 36% (UKB) | 0.61 / 0.85 | **0.60 / 0.76** |
| 69% (GLAD+) | **0.86 / 0.58** | 0.86 / 0.45 |

Bold: values observed in each cohort. p is the prevalence of MDD.

PPV = sensitivity ⨯ p / [ sensitivity ⨯ p + (1- specificity ) ⨯ (1- p)];

NPV = specificity ⨯ (1- p) / [ specificity ⨯ (1- p) + (1-sensitivity) ⨯ p].

**Table S17.** **PPV, NPV, sensitivity and specificity for prediction models in GLAD+ and UK Biobank**

| **Cohort** | **Prediction score cut-off** | **0.3** | **0.4** | **0.5** | **0.6** | **0.7** |
| --- | --- | --- | --- | --- | --- | --- |
| **GLAD+** | PPV | 0.81 | 0.85 | **0.89** | 0.91 | 0.93 |
|  | NPV | 0.67 | 0.62 | **0.58** | 0.53 | 0.47 |
|  | Sensitivity | 0.88 | 0.81 | **0.74** | 0.66 | 0.54 |
|  | Specificity | 0.54 | 0.69 | **0.79** | 0.85 | 0.92 |
| **UKB** | PPV | 0.39 | 0.46 | **0.58** | 0.65 | 0.77 |
|  | NPV | 0.87 | 0.83 | **0.77** | 0.73 | 0.68 |
|  | Sensitivity | 0.96 | 0.85 | **0.60** | 0.45 | 0.22 |
|  | Specificity | 0.15 | 0.43 | **0.75** | 0.86 | 0.96 |

PPV, positive predictive value, true positive / (true positive + false positive).

NPV, negative predictive value, true negative / (true negative + false negative).

Sensitivity, true positive / (true positive + false negative).

Specificity, true negative / (true negative + false positive).

**Table S18. The Pearson’s correlations between predictors in GLAD+ and UK Biobank**

| **Cohort** | **Predictor 1** | **Predictor 2** | **Pearson's correlation** | **95% CI** | **P-value** |
| --- | --- | --- | --- | --- | --- |
| **GLAD+** | Family history of depression | Childhood trauma | 0.17 | 0.15-0.19 | < 2.2e-16 |
|  | Family history of depression | PRS_MDD | 0.15 | 0.13-0.16 | < 2.2e-16 |
|  | Family history of depression | Family history of anxiety | 0.44 | 0.43-0.46 | < 2.2e-16 |
|  | Family history of depression | Sex | 0.18 | 0.17-0.20 | < 2.2e-16 |
|  | Childhood trauma | PRS_MDD | 0.16 | 0.14-0.18 | < 2.2e-16 |
|  | Childhood trauma | Sex | 0.14 | 0.12-0.15 | < 2.2e-16 |
|  | Childhood trauma | Family history of anxiety | 0.13 | 0.11-0.14 | < 2.2e-16 |
|  | Family history of anxiety | PRS_MDD | 0.10 | 0.09-0.12 | < 2.2e-16 |
|  | Family history of anxiety | Sex | 0.14 | 0.12-0.16 | < 2.2e-16 |
|  | PRS_MDD | Sex | 0.08 | 0.06-0.09 | < 2.2e-16 |
| **UKB** | Family history of depression | Childhood trauma | 0.130 | 0.125-0.136 | < 2.2e-16 |
|  | Family history of depression | PRS_MDD | 0.104 | 0.099-0.110 | < 2.2e-16 |
|  | Family history of depression | Family history of anxiety | 0.266 | 0.261-0.272 | < 2.2e-16 |
|  | Family history of depression | Sex | 0.148 | 0.142-0.153 | < 2.2e-16 |
|  | Childhood trauma | PRS_MDD | 0.113 | 0.107-0.119 | < 2.2e-16 |
|  | Childhood trauma | Sex | 0.073 | 0.068-0.079 | < 2.2e-16 |
|  | Childhood trauma | Family history of anxiety | 0.074 | 0.069-0.080 | < 2.2e-16 |
|  | Family history of anxiety | PRS_MDD | 0.067 | 0.061-0.073 | < 2.2e-16 |
|  | Family history of anxiety | Sex | 0.095 | 0.089-0.101 | < 2.2e-16 |
|  | PRS_MDD | Sex | 0.015 | 0.009-0.021 | 5.2e-07 |

**Table S19. Refit the prediction model for MDD to other phenotypes in GLAD+ and UK Biobank**

| **Cohort** | **Dataset** | **Phenotype** | **Beta (SE)** | **R^2^ (%)** | **Scale** | **N** |
| --- | --- | --- | --- | --- | --- | --- |
| **GLAD+** | Training | MDD | 0.54 (0.007) | 32.2 | Liability (K = 5%) | 14,379 |
|  | Testing | MDD episodes | 0.52 (0.008) | 26.8 | Observed | 12,876 |
|  | Testing | MDD age of onset | -0.29 (0.010) | 8.6 | Observed | 9,837 |
|  | Testing | MSM-TRD | 0.07 (0.015) | 0.5 | Observed | 4,206 |
| **UKB** | Training | MDD | 0.41 (0.003) | 23.0 | Liability (K = 15%) | 111,422 |
|  | Testing | MDD episodes | 0.33 (0.003) | 11.1 | Observed | 102,821 |
|  | Testing | MDD age of onset | -0.24 (0.005) | 5.6 | Observed | 38,395 |

MSM-TRD: Maudsley Staging Method (MSM) treatment-resistant depression sum score. The p-value for testing in MSM-TRD is 1.18x10^-05^ in the GLAD+. R² for MDD status is on the liability scale (prevalence assumptions in Table S8); R² for continuous outcomes is on the observed scale and is not directly comparable. ‘Testing’ refers to applying the model trained for MDD status to each severity outcome in the same cohort. K is the prevalence of MDD.

**Table S20.** **The replication of MDD and MDD episodes, age of onset across GLAD+ and UK Biobank**

| **Phenotype** | **Dataset** | **Cohort** | **Beta (SE)** | **R^2^ (%) (95% CI)** | **AUC (95% CI)** | **N** |
| --- | --- | --- | --- | --- | --- | --- |
| **MDD** | Training | GLAD+ | 0.54 (0.007) | 32.2 (30.7-33.7) | 0.83 (0.823-0.837) | 14,379 |
|  | Testing | UKB | 0.41 (0.003) | 23.2 (22.5-23.9) | 0.74 (0.737-0.743) | 111,422 |
|  | Testing | UKB-  resampled | 0.45 (0.018) | 27.9 (23.5-32.3) | 0.78 (0.762-0.798) | 2,442 |
|  | Training | UKB | 0.41 (0.003) | 23.1 (22.4-23.8) | 0.74 (0.737-0.743) | 111,422 |
|  | Testing | GLAD+ | 0.52 (0.007) | 29.8 (28.2-31.4) | 0.82 (0.813-0.827) | 14,379 |
| **MDD episodes** | Training | GLAD+ | 0.51 (0.008) | 26.5 (25.2-27.8) |  | 12,876 |
|  | Testing | UKB | 0.33 (0.003) | 10.1 (9.8-10.4) |  | 102,821 |
|  | Testing | UKB-  resampled | 0.38 (0.019) | 14.5 (11.9-17.1) |  | 2,442 |
|  | Training | UKB | 0.35 (0.003) | 12.1 (11.7-12.5) |  | 102,821 |
|  | Testing | GLAD+ | 0.50 (0.008) | 24.8 (23.5-26.1) |  | 12,876 |
| **MDD age of onset** | Training | GLAD+ | 0.30 (0.010) | 8.8 (7.7-9.9) |  | 9,837 |
|  | Testing | UKB | 0.24 (0.005) | 6.0 (5.5-6.5) |  | 38,395 |
|  | Testing | UKB-  resampled | 0.24 (0.025) | 5.5 (3.3-7.7) |  | 2,442 |
|  | Training | UKB | 0.26 (0.005) | 6.6 (6.1-7.1) |  | 38,395 |
|  | Testing | GLAD+ | 0.29 (0.010) | 8.2 (7.2-9.2) |  | 9,837 |

The UKB resample involves randomly selecting a total of 2,442 (1,597 cases and 845 controls) participants across different episodes, mapping to the distribution of MDD episodes in the GLAD+ study. 95% CIs for AUC were derived using the method of Hanley and McNeil; 95% CIs for R² were derived using the large-sample approximation SE(R²) ≈ √[4R²(1−R²)²/n], where n is the number of MDD cases. Resampled UKB: cases matched to the GLAD+ episode distribution, with randomly selected controls. Sensitivity analysis of case-mix effects; not a comparable cohort. Liability-scale R² assumes K = 5% for GLAD+ and K = 15% for UKB; estimates at common values of K (5-20%) are shown in Table S8.

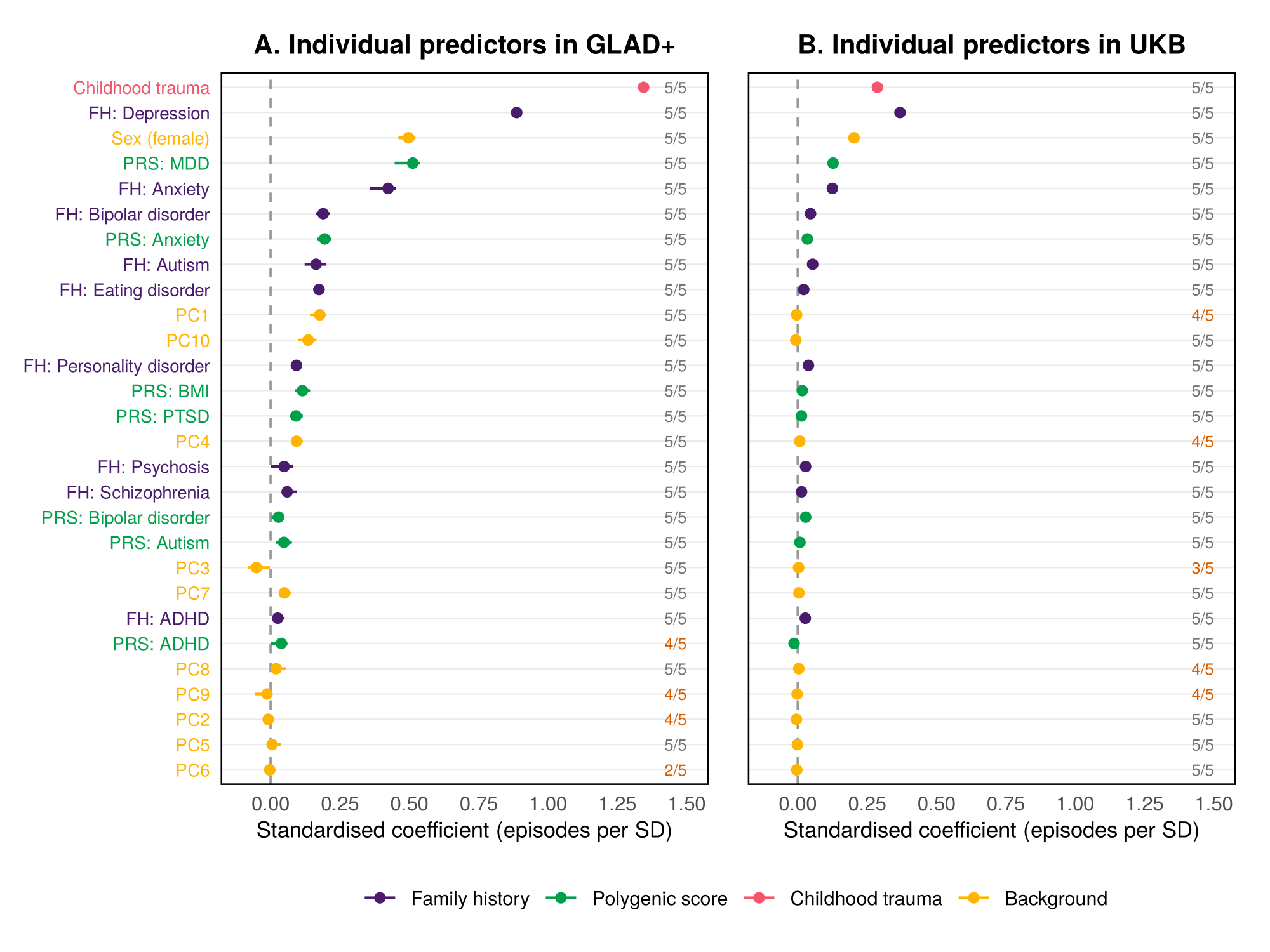

**Figure S3. Contribution of individual predictors to the elastic net model for number of MDD episodes in GLAD+ and UK Biobank**

Standardised coefficients from the elastic net model including all predictors, predicting the number of depressive episodes in GLAD+ (A) and UK Biobank (B). Points show the standardised coefficient (coefficient × predictor SD; change in number of episodes per SD of the predictor), averaged across the five outer-fold models of the nested cross-validation; bars show the range across folds. Labels give the number of outer folds in which the predictor had a non-zero coefficient (orange: fewer than five). Predictors are ordered by their mean absolute standardised coefficient across the two cohorts. Standard deviations of binary predictors were calculated as √[p(1−p)] from their prevalence (Table 1); polygenic scores and genetic principal components were standardised (SD = 1). FH, family history, including family history of 9 psychiatric disorders in GLAD+ and UKB. PRS, polygenic risk scores, including 7 PRSs of psychiatric disorders and BMI based without UK/UKB GWAS sumstats in GLAD+ and UKB. Background, sociodemographic factors, including sex and 10 principal components.

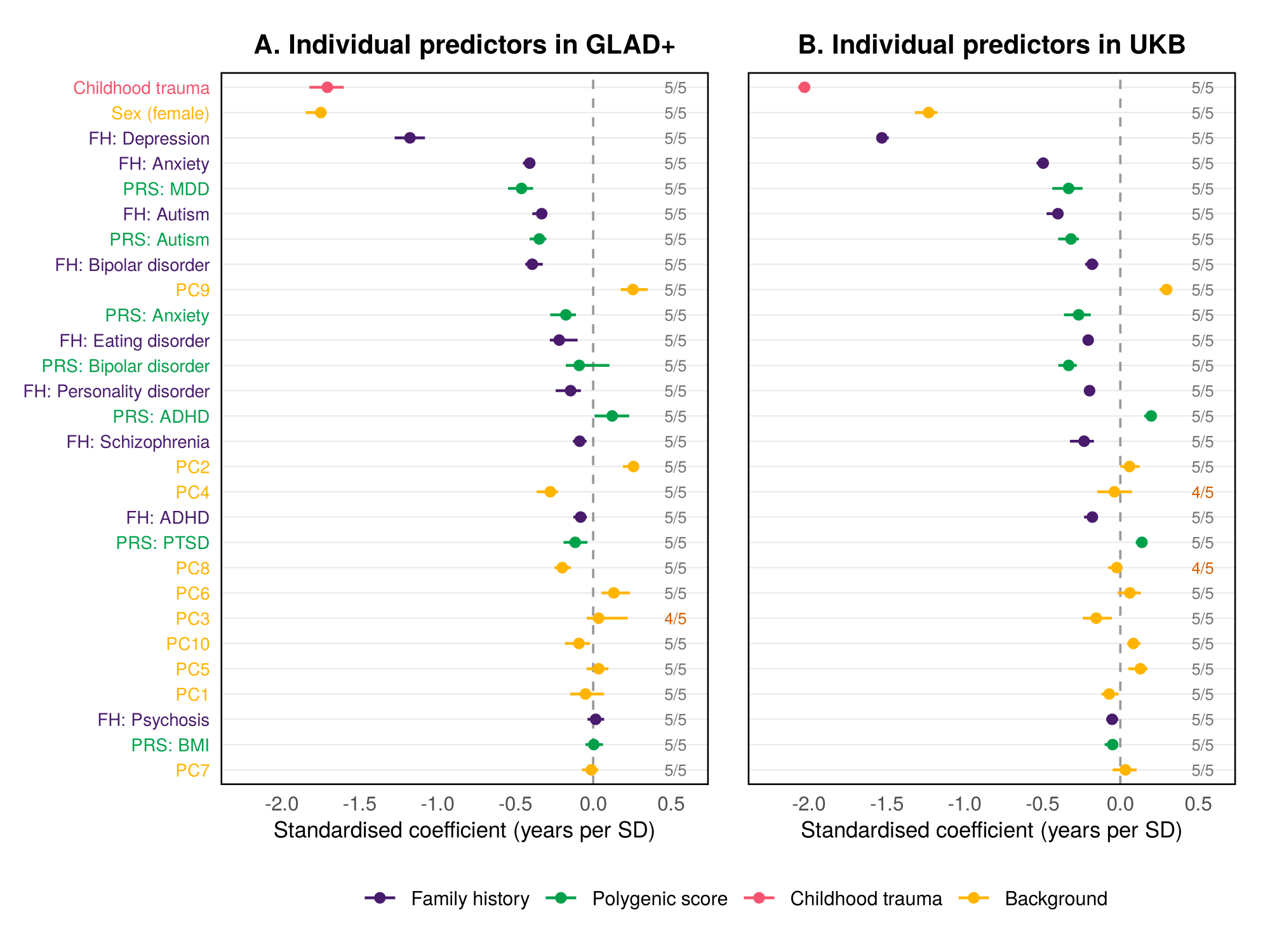

**Figure S4. Contribution of individual predictors to the elastic net model for age of MDD onset in GLAD+ and UK Biobank**

Standardised coefficients from the elastic net model including all predictors, predicting age at onset of MDD in GLAD+ (A) and UK Biobank (B). Points show the standardised coefficient (coefficient × predictor SD; change in age at onset, in years, per SD of the predictor; negative values indicate earlier onset), averaged across the five outer-fold models of the nested cross-validation; bars show the range across folds. Labels give the number of outer folds in which the predictor had a non-zero coefficient (orange: fewer than five). Predictors are ordered by their mean absolute standardised coefficient across the two cohorts. Standard deviations of binary predictors were calculated as √[p(1−p)] from their prevalence (Table 1); polygenic scores and genetic principal components were standardised (SD = 1). FH, family history, including family history of 9 psychiatric disorders in GLAD+ and UKB. PRS, polygenic risk scores, including 7 PRSs of psychiatric disorders and BMI based without UK/UKB GWAS sumstats in GLAD+ and UKB. Background, sociodemographic factors, including sex and 10 principal components.

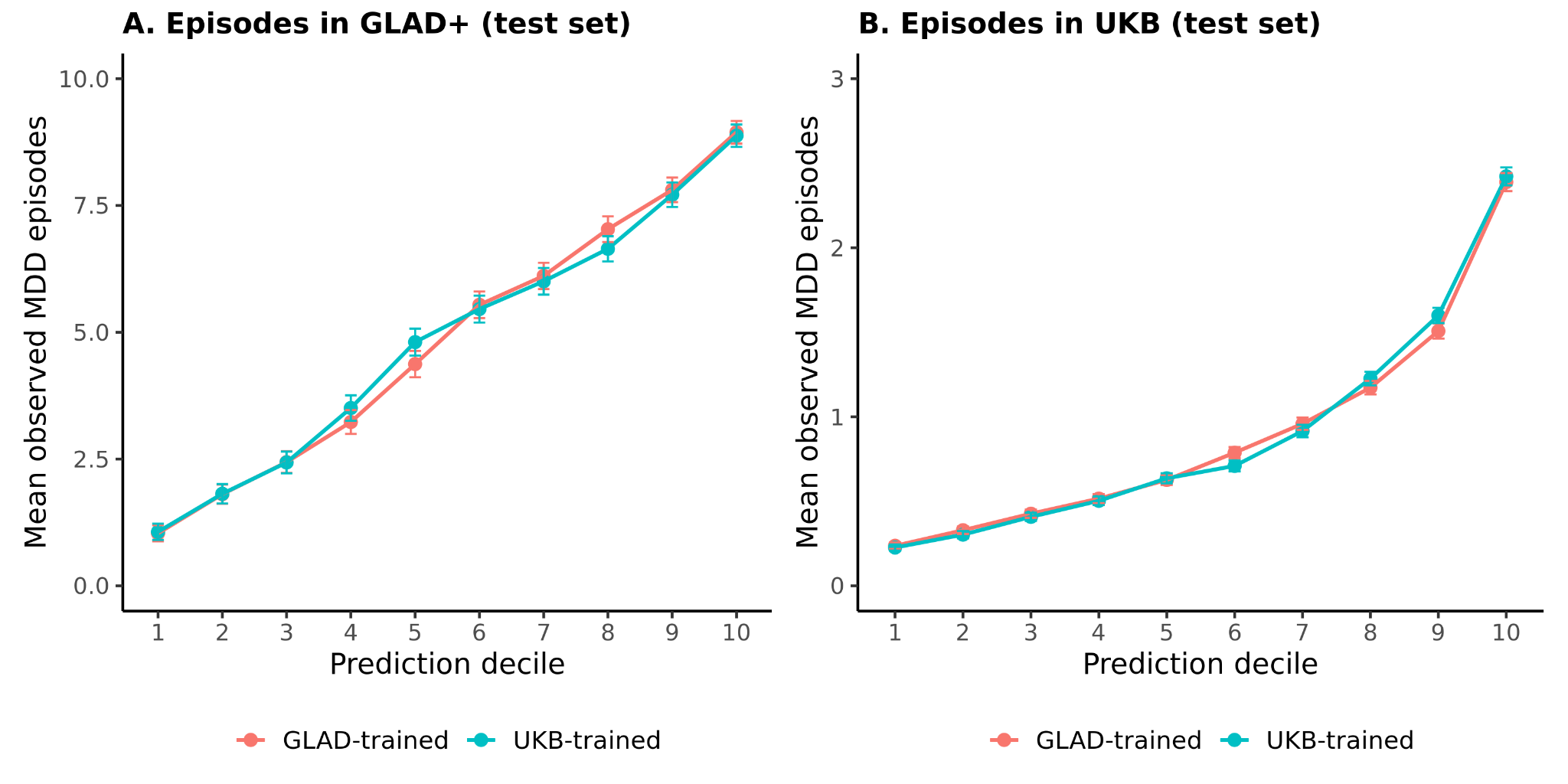

**Figure S5. Decile-based prediction of lifetime MDD episodes in GLAD+ and UK Biobank**

Mean observed lifetime MDD episodes across deciles of predicted risk for GLAD+-trained and UKB-trained models in the GLAD+ test set (A) and UKB test set (B). Both models showed monotonic increases in observed episode counts across prediction deciles in each cohort, supporting consistent relative risk stratification despite differences in absolute scaling between cohorts.

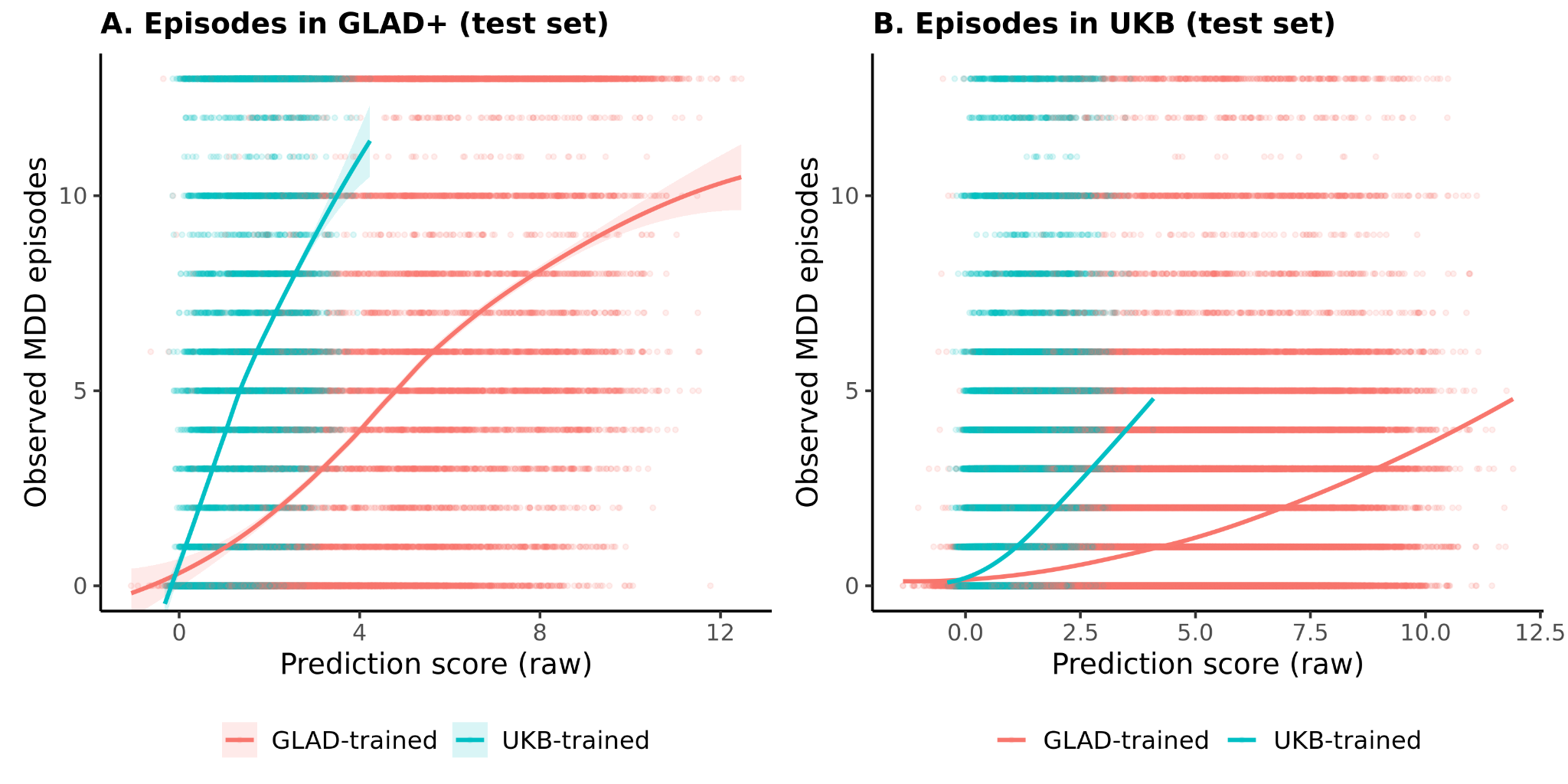

**Figure S6. Raw prediction score against observed lifetime MDD episodes in GLAD+ and UK Biobank**

Observed lifetime major depressive disorder (MDD) episodes plotted against raw prediction scores for models trained in GLAD+ and UK Biobank (UKB). Panel A shows predictions evaluated in the GLAD+ test set, and Panel B shows predictions evaluated in the UKB test set. Points represent individual participants, with horizontal bands reflecting the discrete count nature of the episode measure. Solid lines show smoothed fits of observed episode counts across the raw prediction score for each model, and shaded ribbons indicate 95% confidence intervals. Differences in the slope and range of the curves reflect differences in calibration and dynamic range between within-cohort and cross-cohort prediction.

**Table S21. Software, R packages and versions**

| **Purpose** | **Software / package** | **Version** | **Environment** | **Reference** |
| --- | --- | --- | --- | --- |
| **PRS generation** | | | | |
| PRS pipeline | GenoPred (GenoPredPipe) | January 2024 | HPC | Pain et al., PLoS Genet 2021 |
| PRS method | MegaPRS (BLD-LDAK model) | LDAK v5.1 | HPC | Zhang et al., Nat Commun 2021 |
| Genotype processing | PLINK 1.9 | v1.90b6.10 | HPC | Chang et al., GigaScience 2015 |
| Genotype processing | PLINK 2.0 | v2.00a5.10 | HPC | Chang et al., GigaScience 2015 |
| Genotype imputation (GLAD+) | TOPMed Imputation Server; TOPMed reference panel | Freeze 8 | Web server | Das et al., Nat Genet 2016; Taliun et al., Nature 2021 |
| LD reference | 1000 Genomes Phase 3 (European), HapMap3 variants | – | – | Auton et al., Nature 2015 |
| **Elastic net models** | | | | |
| Statistical environment | R | 4.3.0 | HPC | R Core Team (2023) |
| Elastic net | glmnet | 4.1-8 | R 4.3.0 | Friedman et al., J Stat Softw 2010 |
| Model training and tuning | caret | 7.0-1 | R 4.3.0 | Kuhn, J Stat Softw 2008 |
| Parallel processing | doMC | 1.3.8 | R 4.3.0 | Revolution Analytics |
| Data handling | data.table | 1.17.8 | R 4.3.0 | Barrett et al. |
| Descriptive statistics | psych | 2.4.12 | R 4.3.0 | Revelle |
| Brier score | verification | 1.44 | R 4.3.0 | NCAR |
| ROC and AUC (nested CV) | pROC | 1.18.5 | R 4.3.0 | Robin et al., BMC Bioinformatics 2011 |
| **XGBoost models and AUC comparisons** | | | | |
| Statistical environment | R | 4.4.0 | HPC  /RAP | R Core Team (2024) |
| Gradient boosting | xgboost | 3.1.3.1 | R 4.4.0 | Chen & Guestrin, KDD 2016 |
| DeLong tests (Tables S9, S13) | pROC | 1.19.0.1 | R 4.4.0 | Robin et al., BMC Bioinformatics 2011 |
| **Figures** | | | | |
| Plotting | ggplot2 | 3.5.2 | R 4.4.0 | Wickham, 2016 |

Note: PRS QC, model selection and standardisation are described in the Supplementary Methods (GenoPred pipeline). Random seeds are listed in Table S6.
